# Removing genetic effects on plasma proteins enhances their utility as disease biomarkers

**DOI:** 10.1101/2025.10.14.25337894

**Authors:** Daniela Fusco, Zhijian Yang, Essi Viippola, Tatiana Cajuso, Andrea Corbetta, Chiara Caime, Jakob German, Martina Fu, M. Austin Argentieri, FinnGen, Tomoko Nakanishi, Zhiyu Yang, Andrea Ganna

## Abstract

Plasma protein levels are influenced by both genetic and non-genetic factors and can serve as early disease biomarkers. When a protein is correlated with, but not causally linked to, disease, its genetic determinants can add unwanted variability to protein–disease associations. In such cases, removing the genetic component may improve their predictive performances.

Here, we tested this hypothesis by genetically adjusting 94 highly heritable proteins spanning diverse biological pathways and evaluating their associations with the onset of 37 diseases in 39,871 UK Biobank participants. Genetically adjusted proteins showed stronger associations in 88% of 1,312 significant protein–disease pairs, equivalent to a 30% median reduction in required sample size for comparable power. Of 96 protein–disease pairs with significant differences, all but one showed larger effects for adjusted proteins. Most proteins also showed consistently stronger associations with environmental and lifestyle factors once genetic effects were removed.

Finally, we constructed multi-protein scores from genetically adjusted proteins and demonstrated that they significantly improve prediction for 7 diseases compared to unadjusted proteins. These findings demonstrate that removing genetic effects from plasma proteins is an effective strategy to increase power for biomarker discovery and clinical trial design, consistent with the largely non-causal role of most plasma proteins in disease risk.

## Introduction

Antibody-based, broad-capture proteomics enables the measurement of thousands of plasma proteins across large cohorts^1^. Applied to biobank studies, these scalable approaches have led to the discovery of hundreds of disease biomarkers^2^ and shown that multi-protein scores can strongly predict disease onset across a wide range of disorders, from neurological to cardiometabolic^3,4^.

Plasma protein levels are partially shaped by inherited genetic variation, with the average SNP-based heritability of proteins measured on the Olink Explore platform estimated at ∼0.16^1^. At the same time, proteins capture dynamic, non-genetic risk factors such as smoking, diet, and lifestyle^5–7^, which enhance their value as disease predictors.

Traditionally, genetic effects on protein expression (protein quantitative trait loci, pQTLs) have been leveraged to test causal links between proteins and diseases using Mendelian randomization^8,9^. Here, we take a related but orthogonal approach: removing genetic effects from protein expression to aid biomarker discovery by removing noise due to random genetic variation. This strategy, sometimes referred to as “de-Mendelization”^10^, has not yet been applied at scale, but a notable example is prostate-specific antigen (PSA), where adjusting for genetic determinants of constitutive, non-cancer-related PSA variation improves screening utility^11^.

In this work, we first describe the conditions under which removing genetic effects from proteins can improve the discovery of disease associations. We then demonstrate that, in practice, this strategy consistently increases power to detect protein–disease associations.

## Results

### Conceptual framework

If protein expression causally influences the risk of developing a certain disease, then genetic factors affecting protein expression are also, through the protein, likely to be associated with the disease. This is the fundamental principle underlying Mendelian randomization^12^. In this scenario, removing genetic signals that influence protein expression is expected to weaken the observed association between proteins and disease. This observation extends to the removal of any genetic factor that is causal to the disease (**Figure 1A, upper DAGs**).

**Figure 1:**
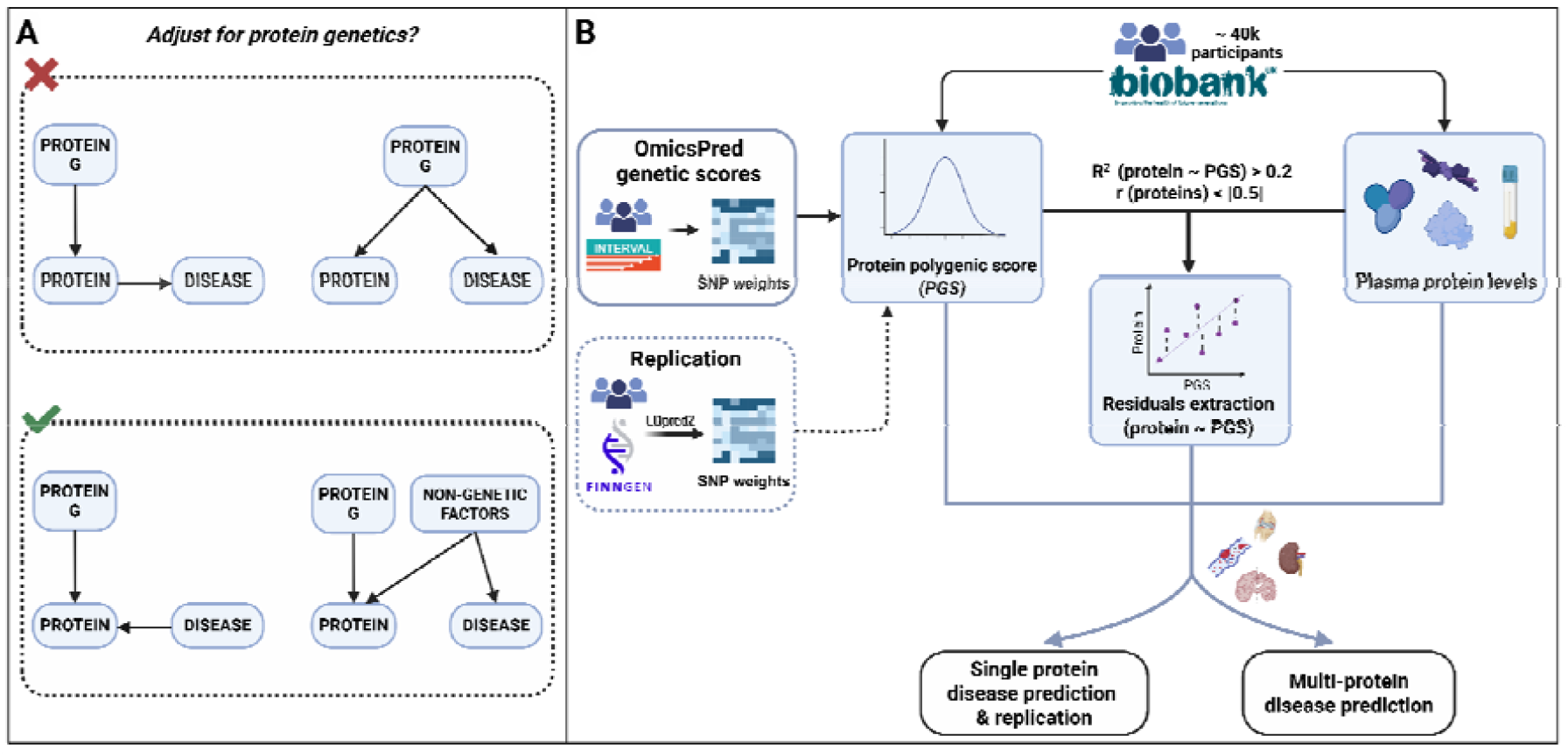
Graphical abstract. **A**. Directed acyclic graphs (DAGs) illustrating how adjusting proteins for their genetically predicted component (G) influences disease prediction. In the top panels (red cross), adjusting for G removes the genetic signal that is causal for disease, thereby reducing the protein’s predictive performance. In contrast (green check), when G is not causal, adjustment may improve prediction. **B**. Protein polygenic scores (PGSs) were derived using OmicsPred genetic scores and calculated in UK Biobank participants of European ancestry. Proteins with high genetic predictability and low correlation were retained. Genetically adjusted proteins were obtained by regressing out the PGS component to extract residuals, which were then used as inputs for both single- and multi-protein disease prediction models. For replication, SNP weights were computed with LDpred2 using FinnGen data. Created with BioRender.com.

However, a protein may not causally influence disease risk. This can occur when protein levels are altered by disease pathophysiology or treatment, or, more commonly, when external non-genetic factors influence both protein levels and disease risk (**Figure 1A, lower DAGs**). Such factors act as confounders of the protein–disease relationship. In this case, genetic influences on proteins represent “unwanted variation” that adds noise to the associations between proteins and disease. In other words, genetics can make proteins less predictive of disease risk by introducing variability unrelated to the disease of interest.

In the latter scenario, removing the genetic effect on a protein can increase the power to detect the association with a disease. In the **Supplementary Methods**, we provide the theoretical derivation showing that the gain in power is a function of both the proportion of protein variance explained by genetics (R^2^) and the strength of the protein–disease association.

### Protein selection and genetic adjustment

We compared protein–disease associations using both directly measured and genetically adjusted protein levels. We initially considered 1,108 proteins measured with Olink or SomaScan in the INTERVAL study^13^, for which OmicsPred provides polygenic score (PGS) weights^14^. These PGSs were calculated in 39,880 participants of European ancestry from the UK Biobank (UKB) (**Figure 1B**). From this set, we retained 94 proteins that were well predicted by their PGS (R^2^ > 0.2) and pruned for correlation (|r| < 0.5) to capture independent biological effects and increase generalizability. Of the 94 proteins, 37 PGSs were derived from genome-wide association studies (GWAS) of Olink-measured proteins and 57 from GWAS of SomaScan-measured proteins. We then regressed the PGS out of each protein, using the residuals as genetically adjusted protein levels (see **Methods**). As a replication analysis, we further considered seven proteins not highly correlated (|r| < 0.5) with the initial 94 and with PGSs weights derived from FinnGen^15^ using an alternative approach.

### Genetically adjusted proteins show stronger associations with disease risk

Among the 94 selected proteins, PGS explained a substantial proportion of variability, with R^2^ values ranging from 20% to 81% (**Supplementary Table 1** and **Supplementary Figure 1**). These proteins span across all the major pathways represented in the overall Olink Explore 3072 panel, including inflammation, neurology, cardiometabolism, and oncogenesis (**Supplementary Table 1**).

We tested associations between the 94 proteins (both unadjusted and genetically adjusted) and the onset of 37 high-burden diseases based on phenotype definitions previously described by Jermy et al.^16^, adjusting for age and sex. This yielded 3,478 protein–disease pairs (**Supplementary Table 2**), of which 1,312 were statistically significant (false discovery rate, FDR < 0.05) in at least one model and were considered in subsequent analyses. All diseases except appendicitis had at least one significant protein association, underscoring the value of plasma proteins as biomarkers across organ systems.

While effect sizes were generally consistent across the two models (**Figure 2A**), genetically adjusted proteins showed an overall trend toward stronger disease associations: in 88% of the 1,312 protein–disease pairs, adjusted proteins had larger absolute z-scores, with an average 57.6% increase in absolute z-scores compared to unadjusted proteins. One way to interpret the improvements in z-scores is to consider the potential gain in statistical power for biomarker discovery, i.e., identifying new protein–disease associations. Among the 1,312 significant protein–disease pairs, the same level of statistical power achieved with unadjusted proteins could have been obtained with a median sample size 30% smaller when using genetically adjusted proteins (**Supplementary Table 3**).

**Figure 2:**
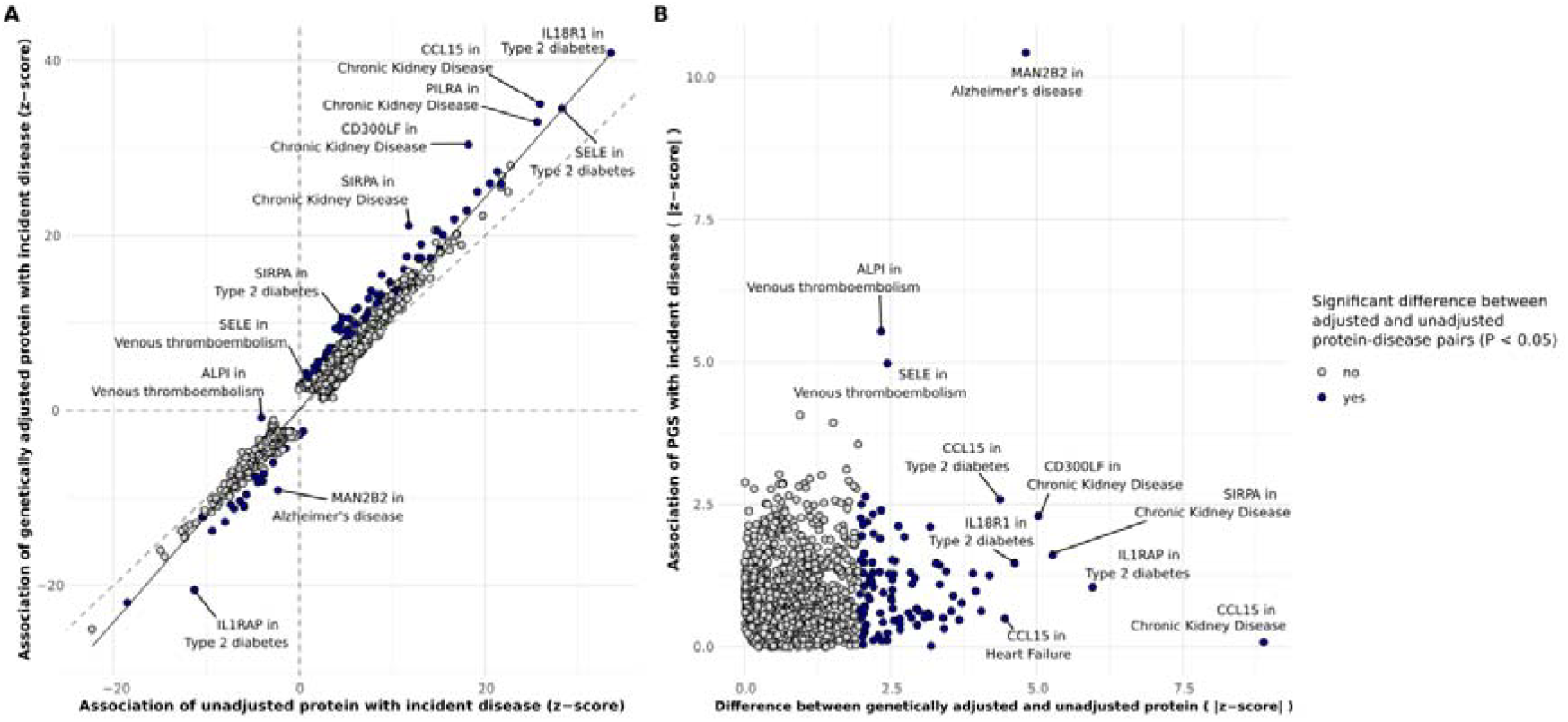
Significant associations between 94 proteins and 37 diseases in UK Biobank. **A**. Logistic regression z-scores for associations with incident diseases for unadjusted proteins (x-axis) and genetically adjusted proteins (y-axis). **B**. Absolute differences in z-score from genetically adjusted and unadjusted protein models (x-axis) and absolute z-score for the association between protein PGS and diseases (y-axis). Blue colour indicates a significant difference in effect sizes between adjusted and unadjusted protein associations (p < 0.05). We report only significant (FDR < 0.05) protein-disease associations in at least one model (unadjusted or adjusted protein); a plot including all pairs is provided as **Supplementary Figure 3**.

We identified 96 protein–disease pairs that showed nominally significantly different effect sizes (p < 0.05) between the genetically adjusted and unadjusted model. Of these, 95 pairs (spanning 43 unique proteins) had stronger effects in the genetically adjusted model, while only one pair showed a stronger effect in the unadjusted model. Type 2 diabetes, chronic kidney disease, and heart failure showed the greatest enrichment (see **Methods**) of associations among the 95 pairs (**Supplementary Figure 2**).

### Stronger disease associations are explained by removal of genetic noise from proteins

To assess whether the observed improvements in the genetically adjusted models can be explained by PGS being strong predictors of protein levels but not directly linked to disease risk, we evaluated associations between protein PGS and disease outcomes with logistic regression (**Figure 2B**).

Among all the 3,478 tested pairs, only four reached significance (FDR < 0.05). As expected, the only protein–disease pair showing a larger effect in the unadjusted model (ALPI and venous thromboembolism) also showed a significant association between ALPI-PGS and venous thromboembolism (P = 3.04 × 10 □□). By contrast, among the 95 protein– disease pairs where genetically adjusted proteins showed stronger effects, most PGS showed weak or no association with disease. Only two pairs reached significance (FDR < 0.05): MAN2B2-PGS with Alzheimer’s disease and SELE-PGS with venous thromboembolism. In these two cases, genetic adjustment does not simply remove “genetic noise” but might induce a spurious association with the disease; we discuss these two cases in more detail in the following sections.

Overall, these results support our hypothesis that the stronger disease associations observed for genetically adjusted proteins are primarily explained by the removal of genetic effects that influence protein variability but do not affect disease risk, thereby reducing noise in protein–disease associations.

### Both cis and trans genetic effects on proteins contribute to stronger disease associations, but cis effects are more robust

The PGS weights calculated by OmicsPred incorporate both cis and trans genetic signals. Among the 94 protein PGS, 55 were constructed using both cis and trans variants, 36 included only cis variants, and 3 contained only trans variants.

For the 55 proteins with both cis and trans scores available, we computed separate cis- and trans-based PGS. Overall, cis-PGS explained more variation in protein expression than trans-PGS. However, for a subset of proteins (e.g., LRPAP1, TNFRSF10C, CD209, FAM3D), trans-PGS showed higher R^2^ (**Supplementary Figure 4**), suggesting that distal genetic regulation also plays a role in the expression of specific proteins.

Then, we repeated the disease association analyses, adjusting proteins separately for cis- or trans-PGS. In the cis-adjusted analysis, among 3,367 protein–disease pairs tested, 1,250 were significantly associated (FDR < 0.05) in either the cis-adjusted or unadjusted models. Seventy-five pairs showed significantly different effect sizes (p < 0.05), all with stronger effects for cis-adjusted proteins (**Supplementary Figure 5**).

A similar but less pronounced pattern was observed for trans-adjusted proteins: among 2,146 pairs tested, 629 were significant at FDR < 0.05, with significantly different effect sizes in 9 pairs—8 stronger for trans-adjusted proteins and only one stronger for unadjusted proteins.

For both cis- and trans-analyses, the improvements in association strength (relative to unadjusted proteins) scaled with PGS prediction accuracy, with the largest gains observed for proteins with higher PGS–protein R^2^ values (**Supplementary Figure 6**).

An important difference between cis and trans analyses was the magnitude of direct associations between protein PGSs and disease. Cis-PGSs, despite being stronger predictors of protein levels, were not associated with any disease (FDR < 0.05). In contrast, trans-PGSs were more likely to associate with diseases, including MAN2B2-transPGS with Alzheimer’s disease and SELE-transPGS with venous thromboembolism (**Supplementary Figure 7**).

These findings indicate that both cis and trans genetic effects can be leveraged to reduce “genetic noise” in proteins and enhance disease prediction, proportionally to how well they genetically predict the protein. However, trans-PGSs, due to the inclusion of a larger number of variants, are more likely to capture pleiotropic variants that are themselves causal for disease. Removing these variants may, in turn, induce spurious correlations between proteins and diseases.

### Illustrative examples of genetically adjusted proteins

We present two examples: one illustrating how genetic adjustment of proteins can improve protein–disease associations, and another showing instead how it may induce spurious associations.

The first example reflects most scenarios where protein-disease associations improved after genetic adjustment. Higher CCL15 protein levels were associated with increased risk of chronic kidney disease (CKD) (OR = 1.68, p = 4.71 × 10□^1^□□; **Figure 3A**), consistent with previous reports showing elevated CCL15 in plasma of patients with chronic renal failure^17^. The PGS for CCL15 was strongly predictive of protein levels (R^2^ = 0.47) but showed no association with CKD (OR = 1.00, p = 0.93). Consequently, genetically adjusted CCL15 levels displayed an even stronger positive association with CKD (OR = 2.20, p = 1.59 × 10□^2^□□). Another way to interpret this result is that the same magnitude of association with CKD observed for unadjusted CCL15 could have been detected with 45% fewer individuals when using genetically adjusted CCL15.

**Figure 3:**
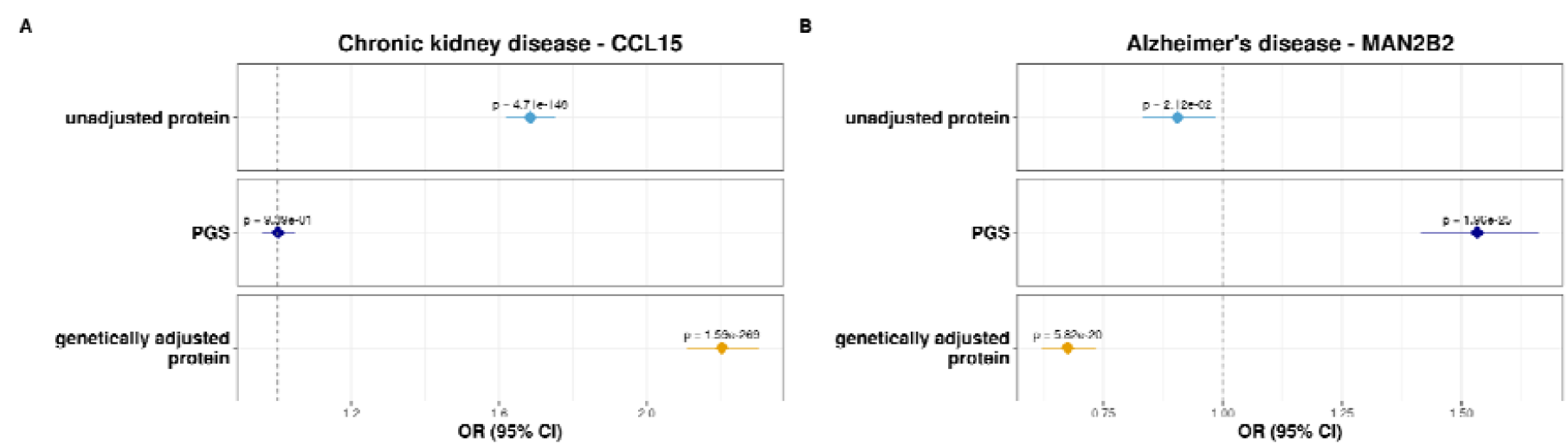
Associations of CCL15 and MAN2B2 with disease risk. Logistic regression Odds Ratios (ORs) with 95% confidence intervals are shown for the association of CCL15 with chronic kidney disease (**A**), and MAN2B2 with Alzheimer’s disease (**B**), using unadjusted protein, PGS, and genetically adjusted protein.

A similar pattern was observed for other CKD-associated proteins, including CD300LF, PILRA, SIRPA, and TDGF1 (**Supplementary Figure 8**), consistent with genetic contributors to these proteins adding noise to the protein–disease association.

A second example highlights a contrasting scenario. Higher MAN2B2 protein levels were weakly associated with decreased risk of Alzheimer’s disease (AD) (OR = 0.91, p = 0.02), whereas higher MAN2B2-PGS was strongly associated with increased AD risk (OR = 1.25, p = 1.06 × 10□^2^□; **Figure 3B**), showing opposite directions of effect. After genetic adjustment, MAN2B2 levels exhibited an even stronger inverse association with AD (OR = 0.86, p = 5.56 × 10 □□).

To investigate this further, we decomposed the PGS into cis and trans components. The association signal was primarily driven by the trans-PGS (OR = 2.02, p = 8.72 × 10□^110^), whereas the cis-PGS showed no significant effect (OR = 0.93, p = 0.072; **Supplementary Figure 9A**). Notably, one of the trans-acting variants included in the PGS mapped to the APOE locus, a well-established genetic risk factor for AD^18^.

These findings suggest that APOE influences both MAN2B2 expression and AD risk through independent mechanisms, supporting a model of horizontal pleiotropy (Figure 1A, upper right DAG) rather than a direct role of MAN2B2 in AD pathogenesis. Consistent with this, there are no prior reports directly implicating MAN2B2 in AD, and protein–protein interaction databases reveal no functional connections between MAN2B2 and APOE.

A similar scenario may underlie the association between genetically adjusted SELE and venous thromboembolism, where the SELE PGS showed a strong association with venous thromboembolism (p = 6.59 × 10□□), only driven by trans effects. Genetic adjustment in this case may induce a spurious association between SELE and venous thromboembolism that is not observed with the unadjusted protein (**Supplementary Figure 9B**)

### Genetically adjusted proteins show stronger correlation with the exposome

By removing genetic effects from proteins, we expect genetically adjusted proteins to exhibit stronger associations not only with diseases but also with non-genetic factors, which we collectively refer to as the exposome. To better characterize which environmental and lifestyle influences are captured by genetically adjusted proteins, we analyzed associations with 86 variables spanning environmental, lifestyle, and sociodemographic domains, along with three laboratory measurements (body mass index (BMI), platelet count, and creatinine). Logistic regressions (or linear regressions for continuous traits) were used to compare associations between the 94 proteins and the exposome, both before and after genetic adjustment (**Figure 4A**). In total, we identified 10,894 significant associations (FDR < 0.05) out of 28,388 protein–exposome pairs tested. Adjusted proteins showed an average 48.7% increase in absolute z-scores compared to unadjusted proteins. Moreover, 921 of 10,894 protein–exposome pairs showed significantly different effect sizes (p < 0.05), and in 96% of these cases, genetically adjusted proteins displayed stronger associations.

**Figure 4:**
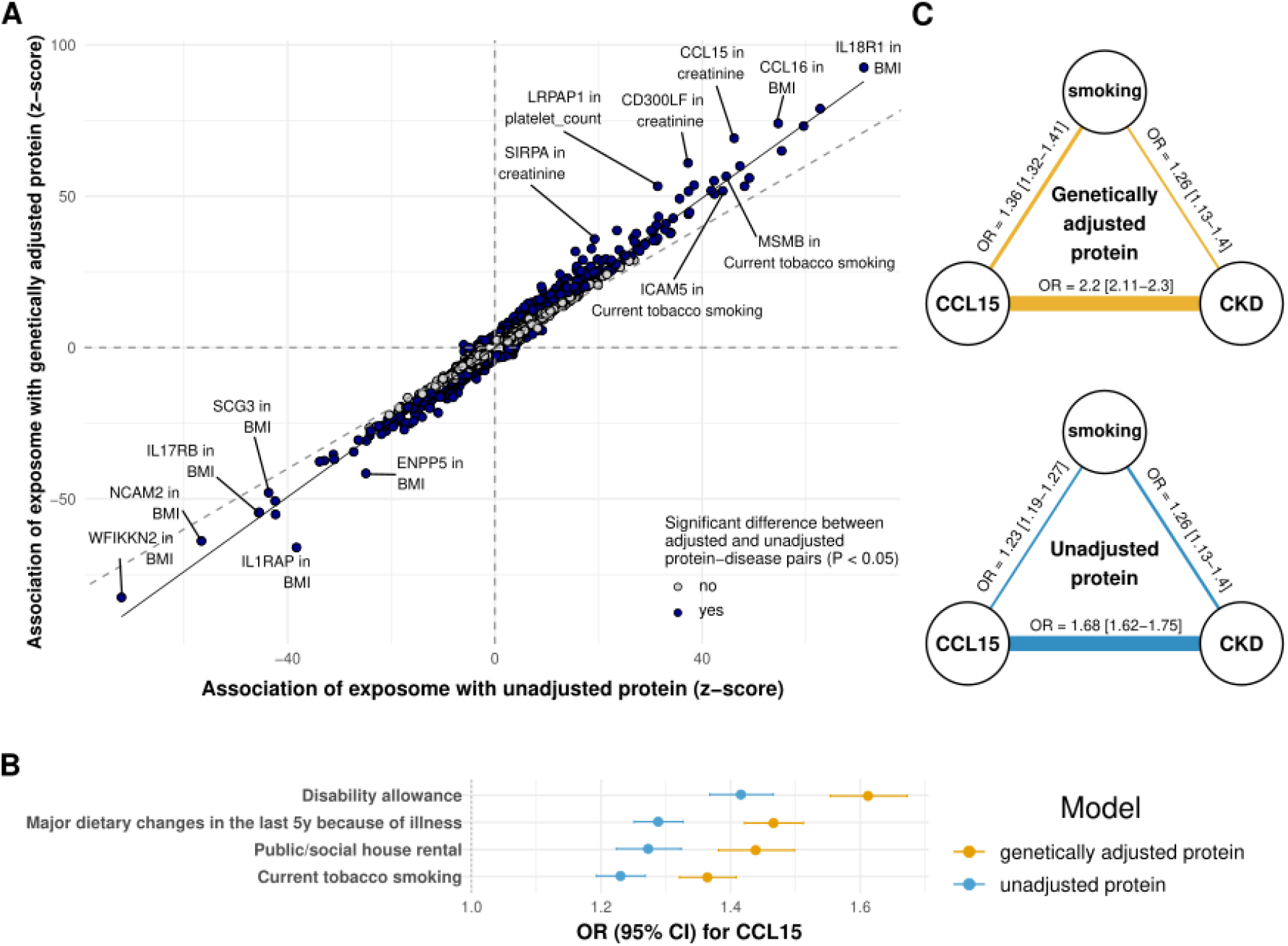
Associations with the exposome. **A**. Associations with exposome features for unadjusted protein (x-axis) versus genetically adjusted protein (y-axis), expressed as z-scores from logistic regressions. Blue colour indicates a significant difference in effect sizes between adjusted and unadjusted protein associations (p < 0.05). We report only significant (FDR < 0.05) protein-disease associations in at least one model (unadjusted or adjusted protein). **B**. Top associated exposures with CCL15, showing OR (95% CI) for the adjusted model (orange) and the unadjusted one (blue). **C**. Associations between CKD and CCL15 unadjusted protein (blue) or adjusted protein (orange), and the “current tobacco smoking” (binarized). 5y= 5 years.

Most association improvements (see **Methods**) were observed for BMI, creatinine, and smoking, consistent with their roles as major factors influencing plasma protein levels. The most enriched proteins included IL1RAP, SIRPA, CD300LF, ENPP5, and CCL15.

Returning to the example of CCL15 and CKD, we found that genetically adjusted CCL15 was more strongly associated with disability benefits, major dietary changes due to illness, and smoking (**Figure 4B**). For instance, the association with current tobacco smoking, a likely confounder of the CCL15–CKD relationship, increased from OR = 1.23 [1.19 – 1.27] to OR = 1.36 [1.32 – 1.41] after genetic adjustment (**Figure 4C**).

Overall, these results suggest that stronger associations between adjusted proteins and the exposome may underlie their improved associations with diseases. In other words, genetically adjusted proteins may serve as better proxies for environmental and lifestyle factors that influence both protein levels and disease risk.

### Multi-protein scores using adjusted proteins are stronger predictors of disease

Previous work has shown that combining multiple proteins can yield powerful predictors of disease risk^3,4^. We compared the standard approach–using protein levels to build multi-protein score–with our approach based on genetically adjusted proteins. Using LASSO regression, we constructed two multi-protein models (unadjusted and genetically adjusted) for the same 37 diseases analyzed previously. Overall, the genetically adjusted models achieved higher AUCs across all diseases, with 7 showing significantly improved performance (one-tailed p < 0.05 for ΔAUC; **Figure 5**).

**Figure 5:**
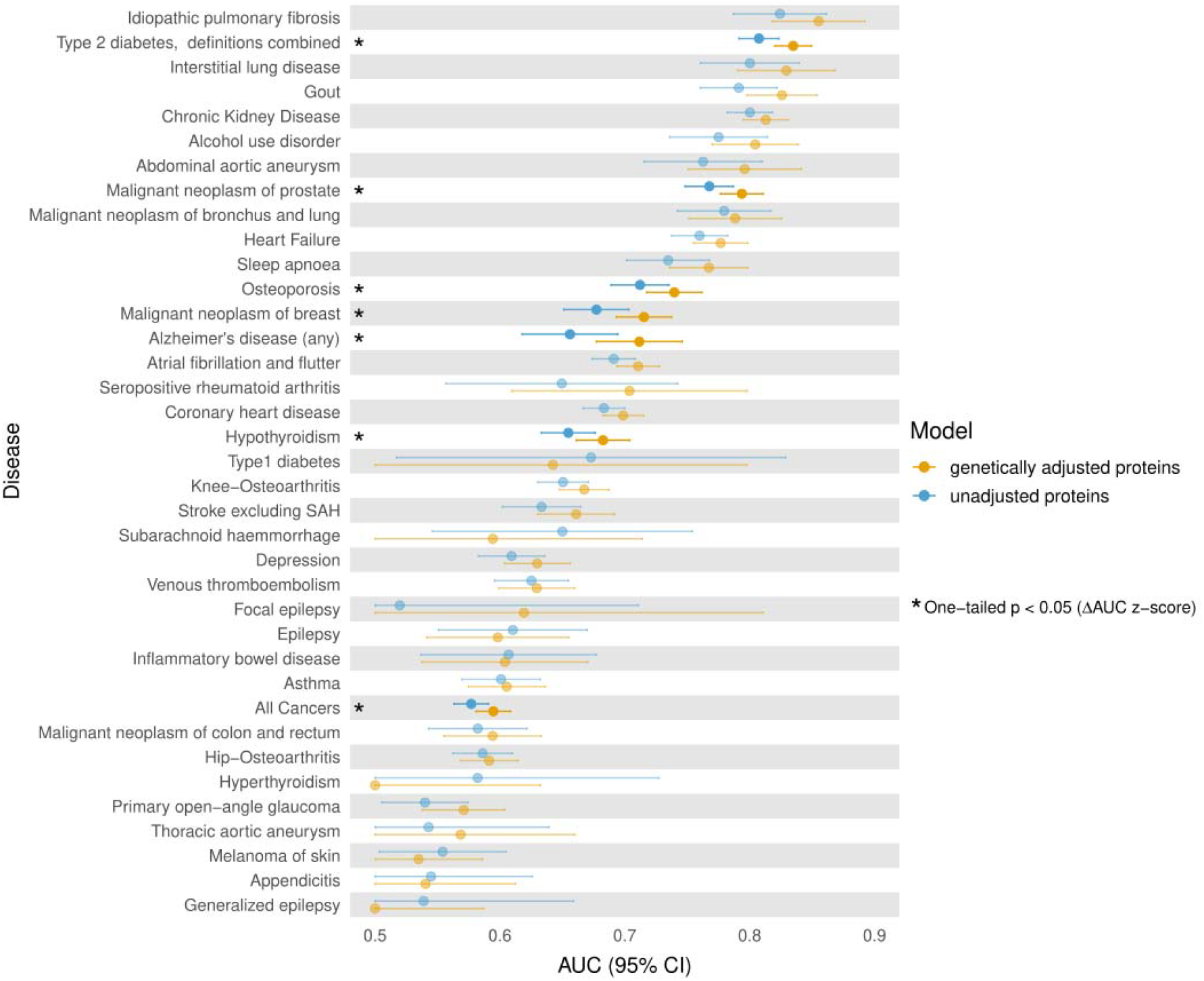
Multi-protein disease prediction. LASSO-derived AUC for disease prediction using unadjusted proteins (blue) or genetically adjusted proteins (orange). Stars and solid-colored points indicate nominal significance (one-tailed p < 0.05) for ΔAUC (AUC_adjusted_-AUC_unadjusted_) z-score.

### Replication using FinnGen-derived polygenic scores and 7 additional proteins

So far, genetic adjustment of proteins was performed using PGS derived by OmicsPred based on GWAS from the INTERVAL study using the SomaScan assay (v.3) and four different panel-specific Olink assays. To further validate our results, we considered protein GWAS from 1,829 individuals in FinnGen measured with the same Olink platform used in the UKB, and we derived PGS using an alternative approach (LDpred2^19^). We identified seven additional proteins not included in the main analysis that met our criteria of low correlation with the existing proteins and strong predictive performance of the PGS. Results were consistent with the main analysis: among 59 significant protein–disease pairs (FDR < 0.05), two showed significantly stronger associations after genetic adjustment, while none favored the unadjusted proteins (**Figure 6**). On average, z-scores increased by 32.1% when using genetically adjusted proteins.

**Figure 6.**
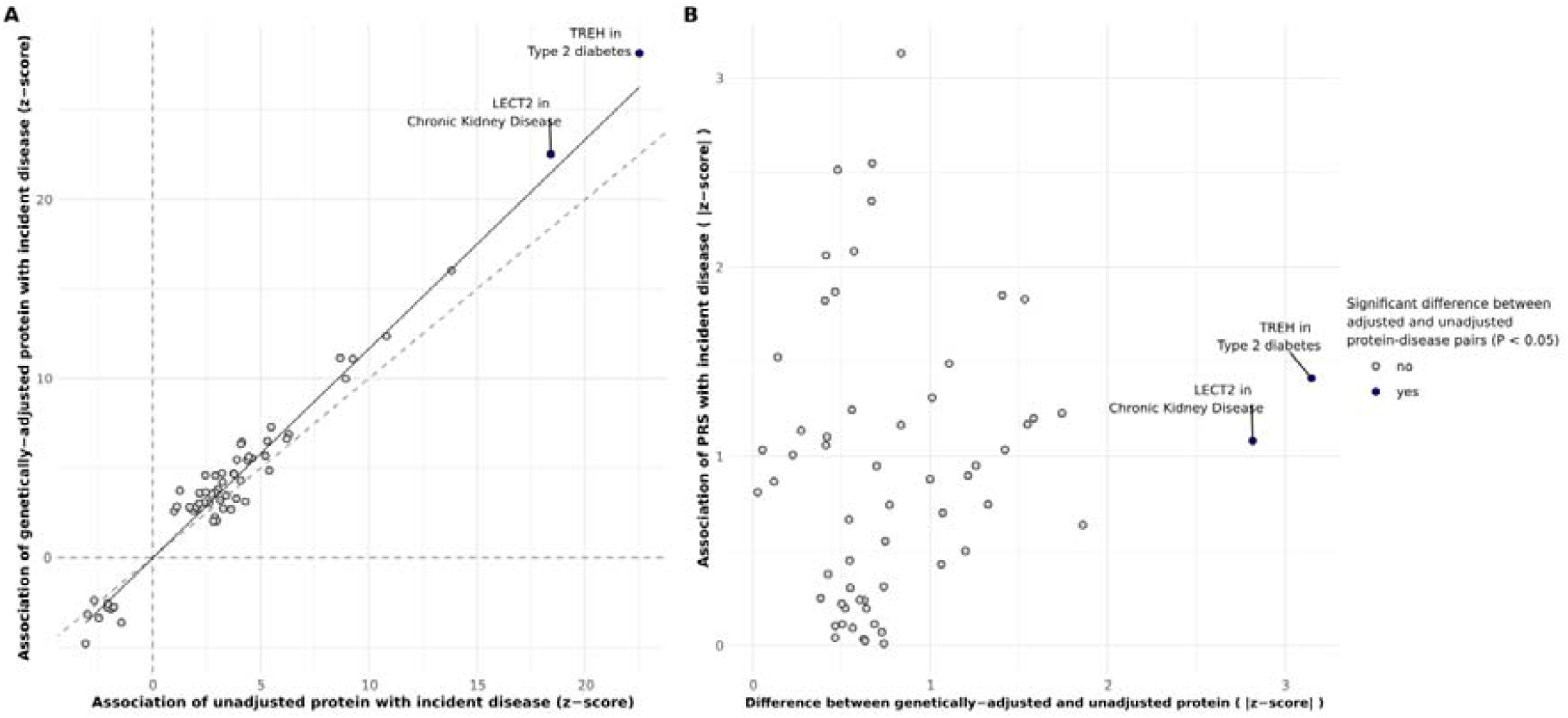
Significant associations between 7 additional proteins and 37 diseases in UK Biobank, using PGS weights from FinnGen. **A**. Logistic regression z-scores for associations with incident diseases for unadjusted proteins (x-axis) and genetically adjusted proteins (y-axis). **B**. Absolute differences in z-score from genetically adjusted and unadjusted proteins models (x-axis) and absolute z-score for the association between protein PGS and diseases (y-axis). Blue colour indicates a significant difference in effect sizes between adjusted and unadjusted protein associations (p < 0.05). We report only significant (FDR < 0.05) protein-disease associations in at least one model (unadjusted or adjusted protein); a plot including all pairs is provided as **Supplementary Figure 10**.

## Discussion

In this study, we show that, in general, removing genetic effects from highly heritable proteins makes them more powerful predictive biomarkers of common disease. This benefit increases with how well PGSs capture protein variation (see **Supplementary Methods** for a technical explanation). Therefore, the value of genetic adjustment is expected to increase as larger protein GWASs are performed and more powerful PGSs are derived.

For most proteins, improvements arise because genetic effects, while being strong predictors of protein levels, are not associated with disease. Removing inter-individual differences in protein levels attributable to inherited genetic variation, therefore, strengthens associations with diseases or environmental factors linked to disease.

Interestingly, genetic adjustment also uncovered associations that were not significant in the unadjusted model. For example, SIRPA showed significant associations with knee osteoarthritis, epilepsy, and three cardiovascular endpoints only after genetic adjustment. SIRPA is best known for its role in the CD47–SIRPA checkpoint^20^, and increased SIRPA expression has been reported in inflamed synovial tissue from rheumatoid arthritis^21^. However, the role of SIRPA as a predictive biomarker for these conditions remains poorly understood and may have been obscured by the strong genetic influence on SIRPA levels (R^2^ = 0.78).

Although it may be tempting to extend the principle of removing “unwanted variation” beyond genetics, it is important to recognize that most non-genetic factors are likely to act as confounders or mediators of the protein–disease relationship, rather than as causal factors for protein levels unrelated to disease. An exception is technical variation (e.g., batch effects), which fits this definition, and adjusting for it has been shown to improve protein– disease associations^22,23^.

A corollary of our findings is that smaller sample sizes are required to detect significant protein–disease associations when using genetically adjusted proteins. We show theoretically (**Supplementary Methods**) that the percentage reduction in required sample size is linearly related to the proportion of protein variance explained by genetics (R^2^). We estimate that the significant protein–disease pairs in our study could have been identified with similar statistical power using, on average, ∼30% fewer individuals. This has implications for the design of biomarker discovery studies and clinical trials where non-causal protein biomarkers are used as outcomes.

To capture the genetic contribution to protein expression, we used polygenic scores (PGSs). While this approach maximizes the proportion of genetic variance explained, it can also introduce biases. In particular, PGS may include variants that influence both protein levels and disease (horizontal pleiotropy), which ideally should not be removed. These pleiotropic effects are more likely to arise from trans variants, simply because they are included in the PGS in larger numbers than cis variants. One illustrative example is MAN2B2 and Alzheimer’s disease: APOE variant has a small trans effect on MAN2B2 but a large causal effect on Alzheimer’s risk. A PGS that includes APOE variants can therefore induce a strong, possibly spurious, association between genetically adjusted MAN2B2 and Alzheimer’s disease. On a technical note, even trans variants with small effects on protein expression can have their impact magnified when combined with strong-effect cis variants in the PGS used for genetic adjustment. In the MAN2B2 example, neither cis-PGS nor trans-PGS adjustment alone induced a spurious association with Alzheimer’s disease, but the combined PGS did (**Supplementary Figure 9A**). We provide a technical explanation of this phenomenon in the **Supplementary Methods**. Based on these observations, and also considering the well-known challenge of PGS transferability across ancestry groups, we suggest using cis-pQTLs rather than genome-wide PGSs when the primary goal is to minimize false positives. Nonetheless, we also note that, overall, trans-adjusted proteins still showed improved disease associations compared to unadjusted proteins.

A limitation of this study is that we considered only 94 proteins in the main analyses and seven in the replication analyses. Whether these findings generalize beyond this set remains to be determined. Of the 1,108 proteins initially evaluated, only 99 had a PGS that satisfied our inclusion criteria (R^2^ > 0.2), and five were excluded due to high correlation with other proteins. The relatively small number of highly genetically predicted proteins can be partly explained by some genetic scores being based on SomaScan and applied to predict Olink proteins in the UK Biobank, where cross-platform correlations between Olink and SomaScan proteins are modest (≈0.3)^23^.

There is no reason to believe that the benefits of genetic adjustment should not extend to proteins with lower PGS R^2^ < 0.2. For our results not to generalize, one would have to assume that highly heritable proteins behave fundamentally differently from less heritable proteins and are less likely to be causal for disease, an assumption for which we find little support. It is instead more likely that most proteins are not causal and instead capture risk factors shared with disease^24^.

Another conceptual limitation is that we did not use genetic information directly for disease prediction. One might argue that, once genetic information is available, it could be added on top of unadjusted proteins to improve disease prediction, rather than being used to remove genetic effects from proteins. While this approach would likely enhance predictive performance, our aim was to demonstrate that genetic adjustment is, in itself, a valuable strategy—particularly when the focus is on protein–disease discovery or when proteins are used as outcomes in intervention studies.

In conclusion, highly heritable proteins are strong predictors of common diseases, but their genetic component often reduces their utility as predictive biomarkers. Accounting for genetic effects offers a promising strategy to enhance biomarker discovery and enable the development of more powerful multi-protein-based prediction models.

## Methods

### Study population

The UK Biobank (UKB) is a prospective cohort of ∼500,000 participants aged 40–69 years at recruitment. We restricted the analyses to individuals of European ancestry (EUR) with available plasma proteomics data, measured using the antibody-based Olink Explore 3072 proximity extension assay^1,25^ (N = 49,005). We restricted our analyses to individuals with at least 2,500 proteins measured at baseline (i.e., the time of recruitment) and whose genetic sex (UKB field ID: 22001) was concordant with their self-reported sex (UKB field ID: 31). After all the filters, the final cohort consisted of 39,880 individuals.

### Proteomics imputation

Within the UKB, we imputed missing values using a random forest-based approach. We first excluded proteins with more than 20% missingness, resulting in a set of 2,919 proteins. We then randomly selected a sample of 10,000 individuals with any genetic ancestry and trained a random forest imputation model using the MissForest^26^ library in Python. We imputed a single dataset using a maximum of five iterations during training, with the other parameters left at default values.

### Selection of OmicsPred genetic scores

We used protein genetic scores from the OmicsPred database^14^, trained on the INTERVAL cohort^13^. We extracted weights derived from both Olink (302 proteins) and SomaLogic (989 proteins) assays, filtering for those also measured in the UKB by the Olink platform. We chose to also include those coming from the SomaLogic assay, given the limited availability of Olink-based summary statistics within the database.Among these proteins, there was an intersection of 183 common proteins, resulting in 1108 unique proteins.

### Polygenic score-based adjustment of proteins

We estimated polygenic score (PGS) for each protein in EUR UKB individuals using PLINK 2.0.0^27^, based on SNP weights obtained from OmicsPred.

Proteins were filtered to retain those with an R^2^ between their observed levels and corresponding PGS exceeded 0.2, using results from the platform with the higher R^2^ in cases where the same protein was included in both platforms. The R^2^ values were calculated using protein measurements at baseline, removing missing values (the sample size varied across proteins).

To reduce redundancy, we further excluded highly correlated proteins (absolute Pearson correlation > 0.5) using the caret::findCorrelation()^28^ function in R, resulting in a final set of 94 proteins.

Then, using previously described imputed values for 39,880 individuals, we isolated the non-genetic component of each single protein by regressing out the variance explained by the corresponding PGS, using linear regression and extracting residuals from it, which were then standardized across individuals. These standardized residuals represent the genetically adjusted proteins used in downstream analyses.

### Disease definitions

We selected 37 diseases based on phenotype definitions previously described by Jermy et al.^16^, extending the panel by applying the same criteria—specifically, harmonization using FinnGen ICD code lists.

We defined “end of follow-up” as the age of the first record of a disease diagnosis (for individuals with the diseases), age at death for a cause other than the disease, age at last record available in the registries or electronic health records, or age 80, whatever occurred first. We excluded prevalent cases, which had their first occurrence before or up to the date of the first assessment visit, and incident cases recorded within the first 6 months of follow-up. A total of 39,871 individuals were included in the analysis. The sample size varied across diseases.

### Disease association analysis

We evaluated associations between proteins and disease endpoints using logistic regression models across all 3,478 protein–disease pairs (94 proteins × 37 diseases). For each pair, three models were fitted independently, each using a different predictor, using the glmnet R package^29^, adjusting for age and sex. We used these predictors: 1) unadjusted protein; 2) genetically adjusted protein; 3) PGS.

All predictors were standardized to facilitate the comparison of effect sizes across models (mean = 0, standard deviation = 1).

Significant associations were defined as those with a false discovery rate (FDR)-adjusted p-value□< □0.05 in either the unadjusted or genetically adjusted protein model.

In order to test for a significant improvement in the associations, we computed the z-score of the difference in effect sizes between the genetically adjusted and the unadjusted protein models, and a two-tailed p-value assessing the significance of the difference.

We performed an enrichment analysis restricted to significant protein–disease pairs. Within this set, we defined a subset of significantly different pairs, in which the absolute z-score was greater for the genetically adjusted model compared to the unadjusted one. For each disease, we counted its occurrences in both the subset and the full set, and tested for over-representation using Fisher’s exact test. Then we ranked diseases by Fisher’s p-value.

To quantify the effective change in statistical power when using unadjusted *vs*. genetically adjusted proteins, we compared the z-scores from the disease-protein association models. The relative sample size R was defined as the squared ratio between the unadjusted and genetically adjusted z-scores. R values greater than 1 indicate that fewer individuals are required to reach the same power when using residualized proteins, and R values smaller than 1 indicate a relative loss of power.

### Cis- trans-analysis

To isolate cis- and trans-acting genetic effects, we defined for each protein the cis-locus as a 1 megabase (Mb) window surrounding the start and end positions of the corresponding gene. To retrieve the genomic positions, we used the GRCh37 Ensembl archive from the biomaRt R package^30^. Using BEDTools^31^, we filtered OmicsPred genetic scores by performing genomic intersection with the defined cis- and trans-loci. The resulting filtered weights were then used to compute PGS following the same methodology described previously. Among the proteins analyzed, three had only trans-pQTLs, 36 had only cis-pQTLs, and 55 had both cis- and trans-pQTLs.

We then extracted the corresponding genetically adjusted proteins from each cis- and trans-PGS, where available. Three proteins had only trans-pQTLs, while 36 proteins had only cis-pQTLs, so 55 proteins had both. Logistic regression analyses were conducted using the cis- and trans-genetically adjusted proteins as predictors for the same disease outcomes, adjusting for age and sex.

### Exposome analysis

To better interpret the information captured by the genetically adjusted proteins, we assessed the association with a set of environmental exposures. We selected a list of exposures with fewer than 1,000 missing values from our dataset of 39,880 individuals, resulting in a final set of 86 traits. In addition, we included three laboratory measurements: body mass index (UKB field ID: 21001), creatinine (UKB field ID: 30700), and platelet count (UKB field ID: 30080).

We performed separate logistic regression analysis for each categorical exposure, and linear regression for each continuous exposure, using the 94 genetically adjusted proteins as predictors and adjusting for age and sex. We further ran a parallel model replacing genetically adjusted proteins with the standardized unadjusted proteins, allowing for a direct comparison of effect sizes. Continuous exposures were standardized prior to analysis.

Categorical exposures were one-hot encoded, with categories represented as separate binary variables; categories with fewer than 100 observations were excluded to ensure model stability. Sample size varied across traits. Significant associations were defined as those with an FDR-adjusted p-value□< □0.05 in either the unadjusted or genetically adjusted protein model.

In order to test for a significant improvement in the associations, we computed the z-score of the difference in effect sizes between the genetically adjusted and the unadjusted protein models, and a two-tailed p-value assessing the significance of the difference.

We performed an enrichment analysis restricted to protein–exposure pairs with FDR < 0.05 in at least one model. Within this set, we defined a subset of pairs with significantly different effect sizes, in which the absolute z-score was greater for the genetically adjusted model. For each exposure, we counted its occurrences in both the subset and the full set of significant pairs, and tested for over-representation using Fisher’s exact test. Then we ranked exposures by Fisher’s p-value. The same procedure was applied to identify the most enriched proteins.

### Multi-protein analysis

To investigate the predictive performance of multiple proteins, we implemented a regularized logistic regression framework using the Least Absolute Shrinkage and Selection Operator (LASSO). As in previous analyses, prevalent cases and incident cases recorded within the first 6 months of follow-up were excluded. For each disease endpoint, we constructed two models using two different sets of predictors: genetically adjusted proteins and unadjusted proteins. For each model, we randomly partitioned the data into training (70%) and test (30%) sets using the caret:createDataPartition()^28^ function in R, matching the proportion of cases and controls in the train and test sets to avoid class imbalance. LASSO logistic regression was fitted to the training data using the glmnet 4.1-9 R package^32^.

We performed 10-fold cross-validation to select the optimal lambda parameter (λ) that minimized binomial deviance (λ.min). Features with non-zero coefficients at the optimal λ were retained as predictive variables. Using the selected features and their coefficients, we computed weighted linear combinations of scores for individuals in both the training and test sets, which were standardized within each set. A secondary logistic regression model was then fitted into the training set using the LASSO-derived score as the predictor. This model was applied to the test set to quantify the predictive performance by calculating the area under the curve (AUC) using the pROC R package^33^. To quantify significant differences, we calculated the z-score of the difference between the AUC of adjusted and unadjusted protein models, and the corresponding one-tailed p-value of this difference.

### Replication with FinnGen pQTLs

We first selected 2,832 proteins from the FinnGen^15^ 3 QCd Olink proteomics dataset of 1,829 independent samples that were also measured in the UKB.

Protein heritability (h^2^) estimation in FinnGen was performed using the Genome-based Restricted Maximum Likelihood (GREML) method, as implemented in GCTA^34^. We first constructed a genetic relationship matrix (GRM) based on 190,180 high-quality, common, and independent SNPs, filtered with the following criteria: INFO score > 0.9, minor allele frequency (MAF) > 0.01, genotyping missingness < 3%, and linkage disequilibrium pruning using a window size of 1,500 kb and pairwise R^2^ < 0.2. GREML analysis was then conducted using the resulting GRM and protein level measurements from 1,829 eligible individuals in the FinnGen cohort.

We then filtered for proteins with a significant h^2^ estimate (p < 0.05) and h^2^ > 0.3, resulting in a set of 237 proteins. To reduce redundancy, we retained proteins with pairwise absolute intercorrelation (Pearson r) below 0.5. Next, we removed proteins that were either present in or highly correlated (absolute Pearson correlation > 0.5 ) with our original set of proteins, yielding a final subset of 135 proteins.

Using FinnGen GWAS data for 135 selected proteins, we applied LDpred2-auto model implemented in the R package bigsnpr to estimate SNP weights for the PGS calculation^19^. We used the HapMap3+ Linkage Disequilibrium (LD) matrix provided with the package^35^, which offers improved genome coverage. After excluding SNPs with minor allele frequencies (MAF) below 1%, a total of 1,301,162 SNPs with available LD information were used. The model failed to converge for two proteins (APOBR and LRRC37A2), which were subsequently excluded. For the remaining 133 proteins, we estimated the PGS for UKB individuals with PLINK 2.0.0 ^27^, based on the SNP weights derived from LDpred2.

Finally, we retained only those proteins (N = 7) for which the variance explained by the PGS (R^2^) in measured protein levels was greater than 0.2, resulting in a high-confidence set of genetically predictable proteins for downstream analysis.

## Data Availability

All data produced in the present study are available upon reasonable request to the authors.

## Acknowledgements

We gratefully acknowledge all participants and research teams involved in UK Biobank and FinnGen.

A.G. has received funding from the European Union’s Horizon 2020 research and innovation program (grant 101016775) and the European Research Council under the same program (grant 945733).

D.F. is a PhD student enrolled in the National PhD in Artificial Intelligence (XXXVIII cycle) course on Health and life sciences, organized by Università Campus Bio-Medico di Roma. We thank Paolo Provero for insightful discussions. We wish to acknowledge CSC – IT Center for Science, Finland, for computational resources.

## Author information

### CreDiT author statement

**Conceptualization**: DF, TN, ZY, AG. **Methodology**: DF, TN, ZY, AG. **Software**: DF, ZhijY, EV, ZY. **Validation**: DF. **Formal analysis**: DF, EV, ZY. **Investigation**: DF, ZhijY, EV, TC, AC, CC, JG, MF, MAA, ZY. **Resources**: FinnGen. **Data curation**: DF, EV. **Writing - original draft**: DF, ZY, AG. **Writing - review & editing**: DF, ZhijY, EV, TC, AC, CC, JG, MF, MMA, TN, ZY, AG. **Visualizations**: DF. **Supervision**: ZY, AG. **Project administration**: DF, AG. **Funding acquisition**: AG.

### Code availability

The code and scripts used in this research will be made available on GitHub (https://github.com/dsgelab/protein-outlier).

### Ethics declarations

This research has been conducted using data from the UK Biobank (http://www.ukbiobank.ac.uk/), a major biomedical database, under approved project ID 78537.

Study subjects in FinnGen provided informed consent for biobank research, based on the Finnish Biobank Act. Alternatively, separate research cohorts, collected prior the Finnish Biobank Act came into effect (in September 2013) and start of FinnGen (August 2017), were collected based on study-specific consents and later transferred to the Finnish biobanks after approval by Fimea (Finnish Medicines Agency), the National Supervisory Authority for Welfare and Health. Recruitment protocols followed the biobank protocols approved by Fimea. The Coordinating Ethics Committee of the Hospital District of Helsinki and Uusimaa (HUS) statement number for the FinnGen study is Nr HUS/990/2017.

The FinnGen study is approved by Finnish Institute for Health and Welfare (permit numbers: THL/2031/6.02.00/2017, THL/1101/5.05.00/2017, THL/341/6.02.00/2018, THL/2222/6.02.00/2018, THL/283/6.02.00/2019, THL/1721/5.05.00/2019 and THL/1524/5.05.00/2020), Digital and population data service agency (permit numbers: VRK43431/2017-3, VRK/6909/2018-3, VRK/4415/2019-3), the Social Insurance Institution (permit numbers: KELA 58/522/2017, KELA 131/522/2018, KELA 70/522/2019, KELA 98/522/2019, KELA 134/522/2019, KELA 138/522/2019, KELA 2/522/2020, KELA 16/522/2020), Findata permit numbers THL/2364/14.02/2020, THL/4055/14.06.00/2020, THL/3433/14.06.00/2020, THL/4432/14.06/2020, THL/5189/14.06/2020, THL/5894/14.06.00/2020, THL/6619/14.06.00/2020, THL/209/14.06.00/2021, THL/688/14.06.00/2021, THL/1284/14.06.00/2021, THL/1965/14.06.00/2021, THL/5546/14.02.00/2020, THL/2658/14.06.00/2021, THL/4235/14.06.00/2021, Statistics Finland (permit numbers: TK-53-1041-17 and TK/143/07.03.00/2020 (earlier TK-53-90-20) TK/1735/07.03.00/2021, TK/3112/07.03.00/2021) and Finnish Registry for Kidney Diseases permission/extract from the meeting minutes on 4th July 2019.

The Biobank Access Decisions for FinnGen samples and data utilized in FinnGen Data Freeze 12 include: THL Biobank BB2017_55, BB2017_111, BB2018_19, BB_2018_34, BB_2018_67, BB2018_71, BB2019_7, BB2019_8, BB2019_26, BB2020_1, BB2021_65,Finnish Red Cross Blood Service Biobank 7.12.2017, Helsinki Biobank HUS/359/2017, HUS/248/2020, HUS/430/2021 §28, §29, HUS/150/2022 §12, §13, §14, §15, §16, §17, §18,§23, §58, §59, HUS/128/2023 §18, Auria Biobank AB17-5154 and amendment #1 (August 17 2020) and amendments BB_2021-0140, BB_2021-0156 (August 26 2021, Feb 2 2022), BB_2021-0169, BB_2021-0179, BB_2021-0161, AB20-5926 and amendment #1 (April 23 2020) and it’s modifications (Sep 22 2021), BB_2022-0262, BB_2022-0256, Biobank Borealis of Northern Finland_2017_1013, 2021_5010, 2021_5010 Amendment, 2021_5018, 2021_5018 Amendment, 2021_5015, 2021_5015 Amendment, 2021_5015 Amendment_2, 2021_5023, 2021_5023 Amendment, 2021_5023 Amendment_2, 2021_5017, 2021_5017 Amendment, 2022_6001, 2022_6001 Amendment, 2022_6006 Amendment, 2022_6006 Amendment, 2022_6006 Amendment_2, BB22-0067, 2022_0262, 2022_0262 Amendment, Biobank of Eastern Finland 1186/2018 and amendment 22§/2020, 53§/2021, 13§/2022, 14§/2022, 15§/2022, 27§/2022, 28§/2022, 29§/2022, 33§/2022, 35§/2022, 36§/2022, 37§/2022, 39§/2022, 7§/2023, 32§/2023, 33§/2023, 34§/2023, 35§/2023, 36§/2023,37§/2023, 38§/2023, 39§/2023, 40§/2023, 41§/2023, Finnish Clinical Biobank Tampere MH0004 and amendments (21.02.2020 & 06.10.2020), BB2021-0140 8§/2021, 9§/2021,§9/2022, §10/2022, §12/2022, 13§/2022, §20/2022, §21/2022, §22/2022, §23/2022, 28§/2022, 29§/2022, 30§/2022, 31§/2022, 32§/2022, 38§/2022, 40§/2022, 42§/2022,1§/2023, Central Finland Biobank 1-2017, BB_2021-0161, BB_2021-0169, BB_2021-0179, BB_2021-0170, BB_2022-0256, BB_2022-0262, BB22-0067, Decision allowing to continue data processing until 31st Aug 2024 for projects: BB_2021-0179, BB22-0067, BB_2022-0262, BB_2021-0170, BB_2021-0164, BB_2021-0161, and BB_2021-0169, and Terveystalo Biobank STB 2018001 and amendment 25th Aug 2020, Finnish Hematological Registry and Clinical Biobank decision 18th June 2021, Arctic biobank P0844: ARC_2021_1001.

### Competing interests

A.G. is the founder of Real World Genetics Oy. The other authors declare no competing interests.

## Notes

### Author Declarations

This research has been conducted using data from the UK Biobank (http://www.ukbiobank.ac.uk/), a major biomedical database, under approved project ID 78537. Study subjects in FinnGen provided informed consent for biobank research, based on the Finnish Biobank Act. Alternatively, separate research cohorts, collected prior the Finnish Biobank Act came into effect (in September 2013) and start of FinnGen (August 2017), were collected based on study-specific consents and later transferred to the Finnish biobanks after approval by Fimea (Finnish Medicines Agency), the National Supervisory Authority for Welfare and Health. Recruitment protocols followed the biobank protocols approved by Fimea. The Coordinating Ethics Committee of the Hospital District of Helsinki and Uusimaa (HUS) statement number for the FinnGen study is Nr HUS/990/2017. The FinnGen study is approved by Finnish Institute for Health and Welfare (permit numbers: THL/2031/6.02.00/2017, THL/1101/5.05.00/2017, THL/341/6.02.00/2018, THL/2222/6.02.00/2018, THL/283/6.02.00/2019, THL/1721/5.05.00/2019 and THL/1524/5.05.00/2020), Digital and population data service agency (permit numbers: VRK43431/2017-3, VRK/6909/2018-3, VRK/4415/2019-3), the Social Insurance Institution (permit numbers: KELA 58/522/2017, KELA 131/522/2018, KELA 70/522/2019, KELA 98/522/2019, KELA 134/522/2019, KELA 138/522/2019, KELA 2/522/2020, KELA 16/522/2020), Findata permit numbers THL/2364/14.02/2020, THL/4055/14.06.00/2020, THL/3433/14.06.00/2020, THL/4432/14.06/2020, THL/5189/14.06/2020, THL/5894/14.06.00/2020, THL/6619/14.06.00/2020, THL/209/14.06.00/2021, THL/688/14.06.00/2021, THL/1284/14.06.00/2021, THL/1965/14.06.00/2021, THL/5546/14.02.00/2020, THL/2658/14.06.00/2021, THL/4235/14.06.00/2021, Statistics Finland (permit numbers: TK-53-1041-17 and TK/143/07.03.00/2020 (earlier TK-53-90-20) TK/1735/07.03.00/2021, TK/3112/07.03.00/2021) and Finnish Registry for Kidney Diseases permission/extract from the meeting minutes on 4th July 2019. The Biobank Access Decisions for FinnGen samples and data utilized in FinnGen Data Freeze 12 include: THL Biobank BB2017_55, BB2017_111, BB2018_19, BB_2018_34, BB_2018_67, BB2018_71, BB2019_7, BB2019_8, BB2019_26, BB2020_1, BB2021_65, Finnish Red Cross Blood Service Biobank 7.12.2017, Helsinki Biobank HUS/359/2017, HUS/248/2020, HUS/430/2021 Sec.28, Sec.29, HUS/150/2022 Sec.12, Sec.13, Sec.14, Sec.15, Sec.16, Sec.17, Sec.18, Sec.23, Sec.58, Sec.59, HUS/128/2023 Sec.18, Auria Biobank AB17-5154 and amendment #1 (August 17 2020) and amendments BB_2021-0140, BB_2021-0156 (August 26 2021, Feb 2 2022), BB_2021-0169, BB_2021-0179, BB_2021-0161, AB20-5926 and amendment #1 (April 23 2020) and its modifications (Sep 22 2021), BB_2022-0262, BB_2022-0256, Biobank Borealis of Northern Finland_2017_1013, 2021_5010, 2021_5010 Amendment, 2021_5018, 2021_5018 Amendment, 2021_5015, 2021_5015 Amendment, 2021_5015 Amendment_2, 2021_5023, 2021_5023 Amendment, 2021_5023 Amendment_2, 2021_5017, 2021_5017 Amendment, 2022_6001, 2022_6001 Amendment, 2022_6006 Amendment, 2022_6006 Amendment, 2022_6006 Amendment_2, BB22-0067, 2022_0262, 2022_0262 Amendment, Biobank of Eastern Finland 1186/2018 and amendment 22Sec./2020, 53Sec./2021, 13Sec./2022, 14Sec./2022, 15Sec./2022, 27Sec./2022, 28Sec./2022, 29Sec./2022, 33Sec./2022, 35Sec./2022, 36Sec./2022, 37Sec./2022, 39Sec./2022, 7Sec./2023, 32Sec./2023, 33Sec./2023, 34Sec./2023, 35Sec./2023, 36Sec./2023, 37Sec./2023, 38Sec./2023, 39Sec./2023, 40Sec./2023, 41Sec./2023, Finnish Clinical Biobank Tampere MH0004 and amendments (21.02.2020 & 06.10.2020), BB2021-0140 8Sec./2021, 9Sec./2021, Sec.9/2022, Sec.10/2022, Sec.12/2022, 13Sec./2022, Sec.20/2022, Sec.21/2022, Sec.22/2022, Sec.23/2022, 28Sec./2022, 29Sec./2022, 30Sec./2022, 31Sec./2022, 32Sec./2022, 38Sec./2022, 40Sec./2022, 42Sec./2022, 1Sec./2023, Central Finland Biobank 1-2017, BB_2021-0161, BB_2021-0169, BB_2021-0179, BB_2021-0170, BB_2022-0256, BB_2022-0262, BB22-0067, Decision allowing to continue data processing until 31st Aug 2024 for projects: BB_2021-0179, BB22-0067, BB_2022-0262, BB_2021-0170, BB_2021-0164, BB_2021-0161, and BB_2021-0169, and Terveystalo Biobank STB 2018001 and amendment 25th Aug 2020, Finnish Hematological Registry and Clinical Biobank decision 18th June 2021, Arctic biobank P0844: ARC_2021_1001.

