## Supplementary Methods for "Removing genetic effects on plasma proteins enhances their utility as disease biomarkers"

Supplementary methods 1: Technical note on how cis- and trans-protein adjustment affects associations

Here, we provide a technical explanation for why even a trans-PRS that is a weak predictor of the protein—but a strong predictor of the disease—can, when considered together with a cis-PRS that is not associated with the disease, lead to a genetically adjusted protein showing a strong and spurious association with the disease. This is what was observed in the case of the MAN2B2–Alzheimer’s disease association.

Let a linear predictor of interest $P$ (the protein) with polygenic determinant $G$ (the PGS) and non-genetic determinants $\epsilon_{P}$(the residuals of the model):

$P=\beta_{PG}G+\epsilon_{P} \#\left( 1 \right)$

Where $G$ and $P$ are standardized ($var\left( G \right)=var\left( P \right)=1)$, $\epsilon_{P}$and $G$ are assumed independent $(\epsilon_{P}\perp G$)

Suppose the polygenic determinant $G$can be divided into two independent genetic components $g_{C}$ and $g_{T}$

$G=\beta_{GC}g_{C}+\beta_{GT}g_{T} \#\left( 2 \right)$

Let $g_{A}$ and $g_{B}$ be homoscedastic ($var(g_{C}) = var(g_{T}) = 1)$, independent between them ($g_{C}\perp g_{T}\to\beta_{GC}^{2}+\beta_{GT}^{2}=1)$, and independent to the non-genetic determinants $\epsilon_{P}$ ($g_{C}\perp\epsilon_{P}$ and $g_{T}\perp\epsilon_{P}$).

*COMMENT: a protein’s PGS can be decomposed into two parts: a pleiotropic (trans) component that also influences the disease and a protein-specific (cis) component that does not. By construction, these components should be approximately independent.”*

Let $g_{T}$ be a causal genetic component (or in LD with the causal genetics) for disease $D$ whereas $g_{C}$ is not:

$D=\beta_{DT}g_{T}+\epsilon_{D} \#\left( 3 \right)$

Where ${var(D) = 1, \epsilon}_{D}$ denotes any other genetic or non-genetic determinants for disease $D$. ${\epsilon_{D}\perp g_{T}, g}_{C}\perp D (therefore {also \epsilon}_{D}\perp g_{C}).$ In this case, a linear unbiased estimate for the association between $D$ and $g_{T}$ would be $\beta_{DT}$, and between $D$ and $g_{C}$ would be 0.

Suppose now we have $\beta_{GC} \gg\beta_{GT}> 0.$

*COMMENT: this corresponds to the association between disease and partial protein PGS separated by pleiotropy. For the MAN2B2 case,* $\beta_{DT}$ *seems to have large magnitude and is greater than 0.*

An unbiased estimate for the association between $D$ and $G$ is:

$\beta_{DG}=\frac{cov\left( D,G \right)}{var\left( G \right)}= Cov\left( \beta_{DT}g_{T}+\epsilon_{D},\beta_{GC}g_{C}+\beta_{GT}g_{T} \right)=\beta_{DT}\beta_{GT} \#\left( 4 \right)$

*COMMENT: this corresponds to the association between disease and full protein PGS. In the MAN2B2 case, this also seems to be rather large, because of large* $\beta_{DT}$*.*

An unbiased estimate for the association between $D$ and $P$ is:

$\beta_{DP}=\frac{cov\left( D,P \right)}{var\left( P \right)}=cov\left( \beta_{DT}g_{T}+\epsilon_{D},\beta_{PG}(\beta_{GC}g_{C}+\beta_{GT}g_{T} \right)+\epsilon_{P})$

$=\beta_{DT}\beta_{PG}\beta_{GT} + cov\left( \epsilon_{D}, \epsilon_{P} \right)\#\left( 5 \right)$

*COMMENT: this corresponds to the association between disease and the unadjusted protein. In the MAN2B2 case, this seems to be very small, because in this case* $cov(\epsilon_{D}, \epsilon_{P})<0$ *(see below), and* $\beta_{DT}\beta_{PS}\beta_{GT} > 0$*.*

If we remove genetic part ($G$) from $P$ (genetic-adjusted protein denoted by $P_{G-}$), an unbiased estimate for the association between $D$ and $P_{G-}$ is:

$\beta_{DP_{G-}}=\frac{Cov\left( D,P_{G-} \right)}{Var\left( P_{G-} \right)}=\frac{cov\left( \beta_{DT}g_{T}+\epsilon_{D},\epsilon_{P} \right)}{var\left( \epsilon_{P} \right)}=\frac{cov\left( \epsilon_{D},\epsilon_{P} \right)}{var\left( \epsilon_{P} \right)}=\frac{cov\left( \epsilon_{D},\epsilon_{P} \right)}{1-\beta_{PG}^{2}} \#\left( 6 \right)$

*COMMENT: this corresponds to the association between disease and the total PGS-adjusted protein. In the MAN2B2 case, this also seems to be negative, indicating* $cov(\epsilon_{D}, \epsilon_{P})<0$*.*

More specifically, if we remove only $g_{C}$ from $G$ (cis-adjusted protein denoted by $P_{C-}$), an unbiased estimate for the association between $D$ and $P_{C-}$ is:

$\beta_{DP_{C-}}=\frac{Cov\left( D,P_{C-} \right)}{Var\left( P_{C-} \right)}=\frac{cov\left( \beta_{DT}g_{T}+\epsilon_{D},{\beta_{PG}\beta_{GT}g_{T} +\epsilon}_{P} \right)}{var\left( {\beta_{PG}\beta_{GT}g_{T} +\epsilon}_{P} \right)} =\frac{{\beta_{DT}\beta}_{PG}\beta_{GT}+ cov\left( \epsilon_{D},\epsilon_{P} \right)}{var\left( {\beta_{PG}\beta_{GT}g_{T} +\epsilon}_{P} \right)} = \frac{{\beta_{DT}\beta}_{PG}\beta_{GT}+ cov\left( \epsilon_{D},\epsilon_{P} \right)}{1-\left( \beta_{PG}\beta_{GC} \right)^{2}} \#\left( 7 \right)$

*COMMENT: this corresponds to the association between disease and the “cis”-adjusted protein. In the MAN2B2 case, this also seems to be very small. Note that the numerator in this form is the same as the disease association with the unadjusted protein, which is almost 0 as observed above.*

if we remove only $g_{B}$ from $P$ (cis-adjusted protein denoted by $P_{B-}$), an unbiased estimate for the association between $D$ and $P_{B-}$ is:

$\beta_{DP_{T-}}=\frac{Cov\left( D,P_{T-} \right)}{Var\left( P_{T-} \right)}$

$=\frac{cov\left( \beta_{DT}g_{T}+\epsilon_{D},{\beta_{PG}\beta_{GC}g_{C} +\epsilon}_{P} \right)}{var\left( {\beta_{PG}\beta_{GC}g_{C} +\epsilon}_{P} \right)}$

$=\frac{cov\left( \epsilon_{D},\epsilon_{P} \right)}{var\left( {\beta_{PG}\beta_{GC}g_{C} +\epsilon}_{P} \right)} = \frac{cov\left( \epsilon_{D},\epsilon_{P} \right)}{1-\left( \beta_{PG}\beta_{GT} \right)^{2}} \#\left( 8 \right)$

*COMMENT: this corresponds to the association between disease and the “trans”-adjusted protein. In the MAN2B2 case, this also seems to be very small.*

Note the numerator of $\beta_{DP_{T-}}$(disease association with the trans-adjusted protein) in (8) is the same as $\beta_{DP_{G-}}$ (disease association with the PGS-adjusted protein) in (6) when the hypothesis in (3) holds. However, we see the association $\beta_{DP_{T-}}$ smaller than $\beta_{DP_{G-}}$because their denominators $1-\left( \beta_{PG}\beta_{GT} \right)^{2} >>1-\beta_{PG}^{2}$ (due to small $\beta_{GT}$). Therefore:

$|\frac{cov\left( \epsilon_{D},\epsilon_{P} \right)}{1-\beta_{PG}^{2}}|>|\frac{cov\left( \epsilon_{D},\epsilon_{P} \right)}{1-\left( \beta_{PG}\beta_{GT} \right)^{2}}|\to|\beta_{DP_{G-}}|>| \beta_{DP_{T-}} |\#\left( 9 \right)$

CONCLUSION

Small-effect trans variants can be amplified during genetic adjustment when combined with strong cis effects in the PGS.

Supplementary Methods 2: Relationship between power gain and proportion of protein variance explained by genetics

Here, we provide a technical explanation of why, when protein genetics is not associated with the disease of interest, if genetics explains much of a protein’s variability, then removing that genetic part from the protein boosts the statistical power to detect its association with disease.

Let a linear predictor of interest $P$ (the protein) with polygenic determinant $G$ (the PGS) and non-genetic determinants $\epsilon_{P}$ (the residuals of the model):

$P=\beta_{PG}G+\epsilon_{P}\#\left( 1 \right)$

Where G and P are standardized ($var\left( G \right)=var\left( P \right)=1)$, $\epsilon_{P}$and $G$ are assumed independent $(\epsilon_{P}\perp G$). Since $P$ depends on $G$ with coefficient $\beta_{PG}$, the proportion of variance of $P$ explained by $G$ is $\beta_{PG}^{2}$ and the residual variance of P is then $var\left( \epsilon_{P} \right)$ = ${1-\beta}_{PG}^{2}$.

Let a target disease $D$ with $var\left( D \right)=1$, suppose the genetics of $P$ is independent of $D$ ($D\perp G$).

The unbiased estimate for the linear regression coefficient of $D$ on $P$ is:

$\hat{\beta_{DP}}=\frac{cov\left( P,D \right)}{var\left( P \right)}=\frac{cov\left( \beta_{PG}G+\epsilon_{P},D \right)}{1}=\beta_{PG} cov\left( G,D \right)+cov\left( \epsilon_{P},D \right)=cov\left( \epsilon_{P},D \right)\#\left( 2 \right)$

By the definition of the variance of an ordinary least squares (OLS) estimate, it can be written as:

$var\left( \hat{\beta_{DP}} \right)=\frac{var\left( D-\hat{\beta_{DP}}P \right)}{\left( n-1 \right)var\left( P \right)}=\frac{var\left( D-\hat{\beta_{DP}}P \right)}{n-1}=$

$\frac{var\left( D \right)+{\hat{\beta_{DP}}}^{2}var\left( P \right)-2\hat{\beta_{DP}}cov\left( \epsilon_{P},D \right)}{n-1}=\frac{1+{\hat{\beta_{DP}}}^{2}-2{\hat{\beta_{DP}}}^{2}}{n-1}=\frac{1-{\hat{\beta_{DP}}}^{2}}{n-1}\#\left( 3 \right)$

Therefore

$z_{DP}=\frac{\hat{\beta_{DP}}}{SE_{\hat{\beta_{DP}}}}=\frac{\hat{\beta_{DP}}}{\sqrt{\frac{1-{\hat{\beta_{DP}}}^{2}}{n-1}}} \#\left( 4 \right)$

Now, let us remove genetics $G$ from $P$ hence evaluating only $\epsilon_{P}$ (in our case the residuals of the linear model), the unbiased estimate for the linear regression coefficient of $P_{G-} on D$ becomes:

$\hat{\beta_{DP_{G-}}}=\frac{cov\left( \epsilon_{P},D \right)}{var\left( \epsilon_{P} \right)}=\frac{cov\left( \epsilon_{P},D \right)}{{1-\beta}_{PG}^{2}}=\frac{\hat{\beta_{DP}}}{{1-\beta}_{PG}^{2}} \#\left( 5 \right)$

With a variance of

$var\left( \hat{\beta_{DP_{G-}}} \right)=\frac{var\left( D-\hat{\beta_{DP_{-S}}\epsilon_{P}} \right)}{\left( n-1 \right)var\left( \epsilon_{P} \right)}=\frac{var\left( D-\frac{\hat{\beta_{DP}}}{{1-\beta}_{PG}^{2}}\epsilon_{P} \right)}{\left( n-1 \right)\left( {1-\beta}_{PG}^{2} \right)}$

$=\frac{var\left( D \right)+\left( \frac{\hat{\beta_{DP}}}{{1-\beta}_{PG}^{2}} \right)^{2}var\left( \epsilon_{P} \right)-\frac{2\hat{\beta_{DP}}}{{1-\beta}_{PG}^{2}}cov\left( \epsilon_{P},D \right)}{\left( n-1 \right)\left( {1-\beta}_{PG}^{2} \right)}$

$=\frac{1+\left( \frac{\hat{\beta_{DP}}}{{1-\beta}_{PG}^{2}} \right)^{2}\left( {1-\beta}_{PG}^{2} \right)-\frac{2\hat{\beta_{DP}^{2}}}{{1-\beta}_{PG}^{2}}}{\left( n-1 \right)\left( {1-\beta}_{PG}^{2} \right)}=\frac{\frac{{1-\beta}_{PG}^{2}-\hat{\beta_{DP}^{2}}}{1-\beta_{PG}^{2}}}{\left( n-1 \right)\left( {1-\beta}_{PG}^{2} \right)}=\frac{{1-\beta}_{PG}^{2}-\hat{\beta_{DP}^{2}}}{\left( n-1 \right)\left( {1-\beta}_{PG}^{2} \right)^{2}}\#\left( 6 \right)$

and a z-score of:

$z_{DP_{G-}}=\frac{\hat{\beta_{DP_{G-}}}}{SE_{\hat{\beta_{DP_{G-}}}}}=\frac{\frac{\hat{\beta_{DP}}}{{1-\beta}_{PG}^{2}}}{\sqrt{\frac{{1-\beta}_{PG}^{2}-\hat{\beta_{DP}^{2}}}{\left( n-1 \right)\left( {1-\beta}_{PG}^{2} \right)^{2}}}}=\frac{\hat{\beta_{DP}}}{\sqrt{\frac{{1-\beta}_{PG}^{2}-\hat{\beta_{DP}^{2}}}{n-1}}}\#\left( 7 \right)$

Let us compare the z-score obtained with their ratio:

$\frac{z_{DP_{G-}}}{z_{DP}}=\frac{\frac{\hat{\beta_{DP}}}{\sqrt{\frac{{1-\beta}_{PG}^{2}-\hat{\beta_{DP}^{2}}}{n-1}}}}{\frac{\hat{\beta_{DP}}}{\sqrt{\frac{1-{\hat{\beta_{DP}}}^{2}}{n-1}}}}=\frac{\sqrt{1-{\hat{\beta_{DP}}}^{2}}}{\sqrt{{1-\beta}_{PG}^{2}-\hat{\beta_{DP}^{2}}}}\#\left( 8 \right)$

This means that the more variance the polygenic score explains in the protein ($\beta_{PS}^{2}$, i.e. the $R^{2}$), the larger the ratio of $z$-scores becomes when we regress the disease on the non-genetic component of the protein (residualised on genetics) versus the unadjusted protein. In other words, when genetics of P is not associated with target disease D, the stronger the genetic contribution to $P$, the more power we gain by removing that genetic component when testing the association with disease.

Note that since by the definition of Z-score:

$Z=\frac{\underline{x}-\mu}{\frac{\sigma}{\sqrt{n}}}$

Where $\underline{x}$ and $\mu$ are sample and population mean respectively and $\sigma$ is the population standard deviation, a ratio of squared z-scores can also be interpreted as the *reciprocal* of required sample size ($n$) ratio for detecting a same effect. Therefore, based on $\#\left( 8 \right)$, to detect a same association effect, comparing to $P$, using $P_{G-}$ may reduce the sample size requirement by:

$1-\left( \frac{z_{DP}}{z_{DP_{G-}}} \right)^{2}=1-\frac{{1-\beta}_{PG}^{2}-\hat{\beta_{DP}^{2}}}{1-{\hat{\beta_{DP}}}^{2}}=\frac{\hat{\beta_{PG}^{2}}}{1-{\hat{\beta_{DP}}}^{2}}$

CONCLUSION

The percentage reduction in required sample size is linearly related to the proportion of protein variance explained by genetics (R² or $\beta_{PG}^{2}$)
