## Supplementary Figures for "Removing genetic effects on plasma proteins enhances their utility as disease biomarkers"

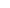

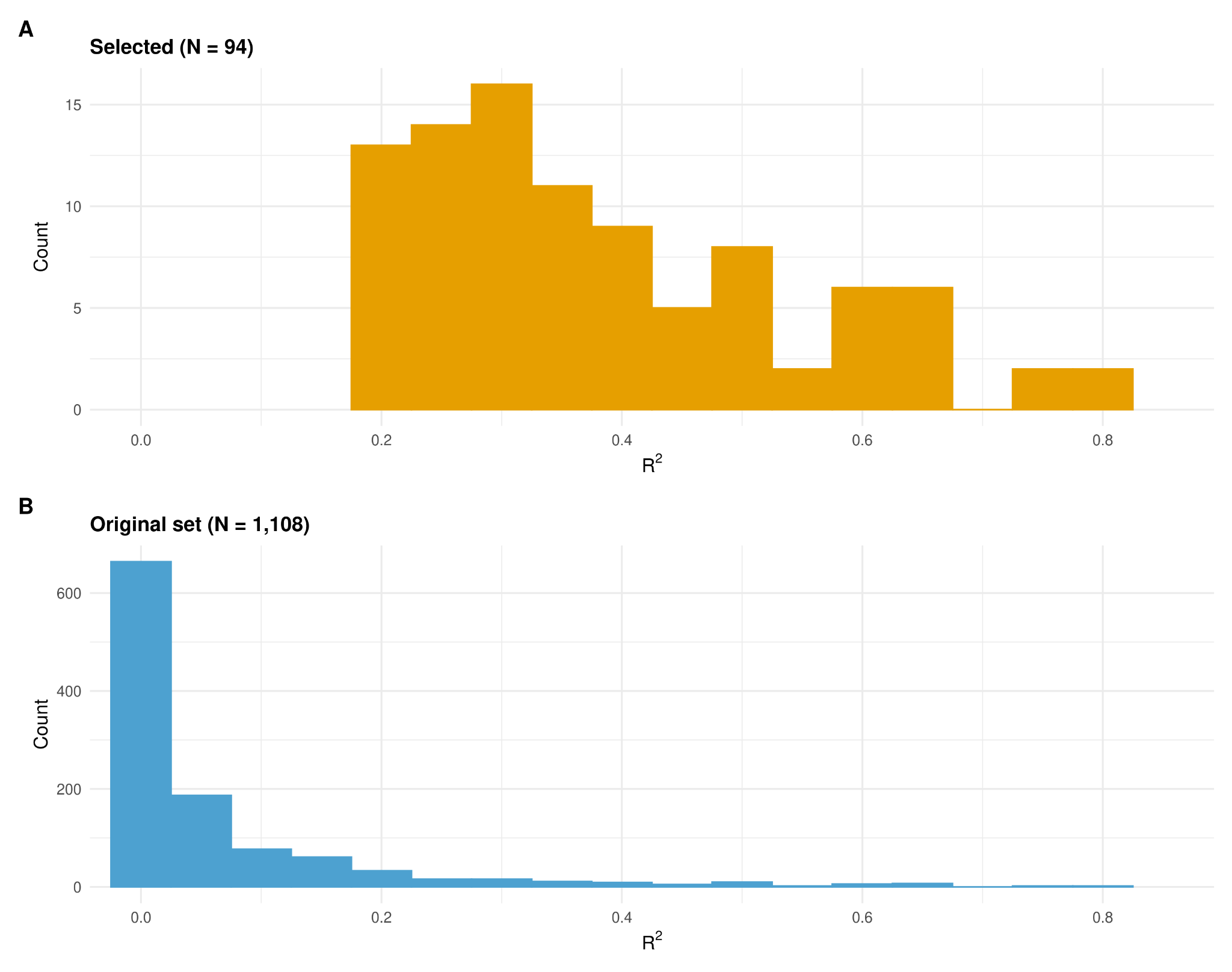

**Supplementary Figure 1 Protein variance explained by PGS (R^2^).** Distributions of the R^2^ between protein levels and PGS across the 94 selected proteins (A) and the original set of proteins before the filters (B).

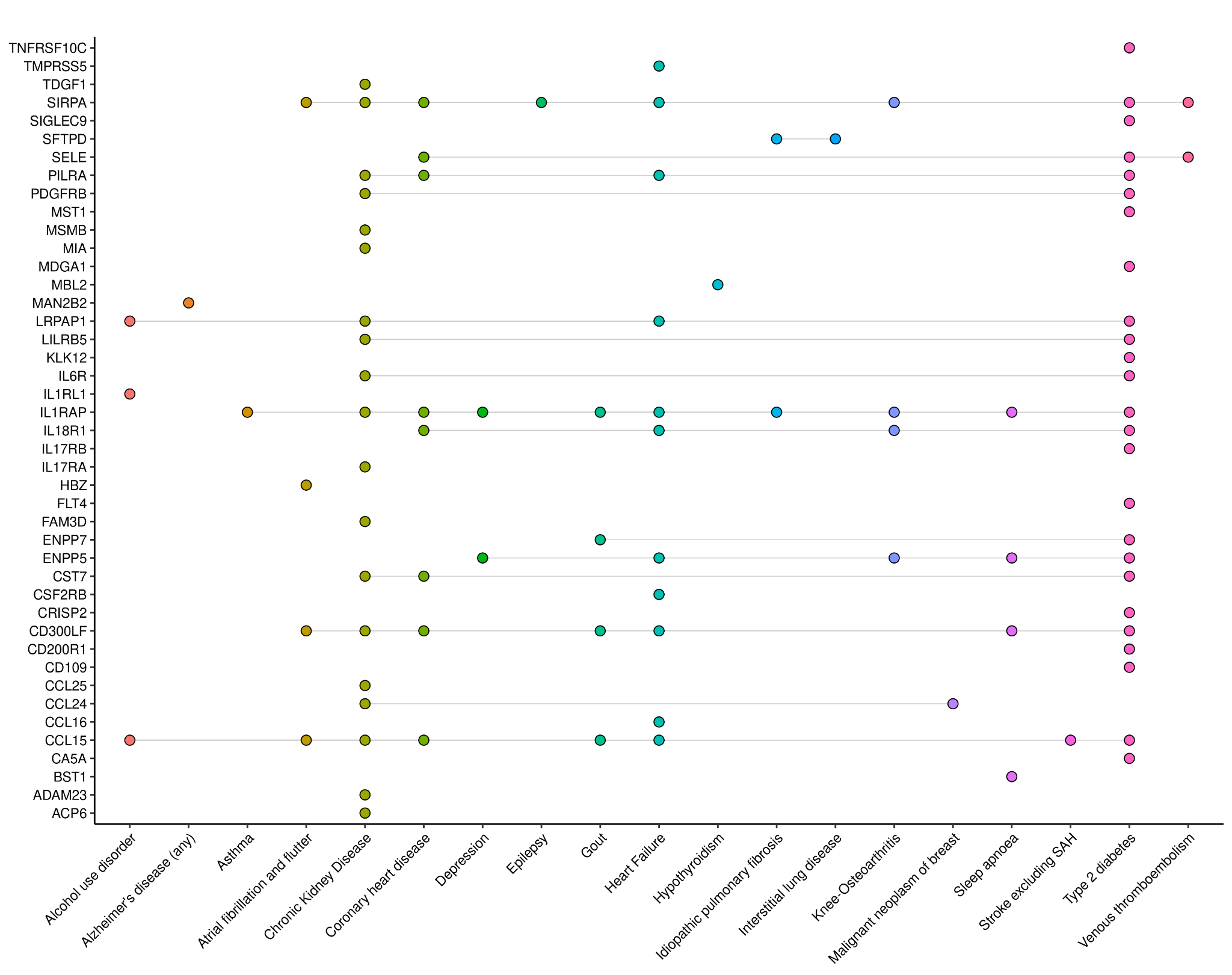

**Supplementary Figure 2 Upset plot of the top 95 protein-disease pairs showing stronger effects in the genetically adjusted vs unadjusted analysis.** Dots represent a significant association (FDR < 0.05 in at least one model and p of the difference < 0.05) between a disease (x-axis) and a protein (y-axis). Gray lines indicate multiple diseases associated with the same protein, and dot colours represent each disease.

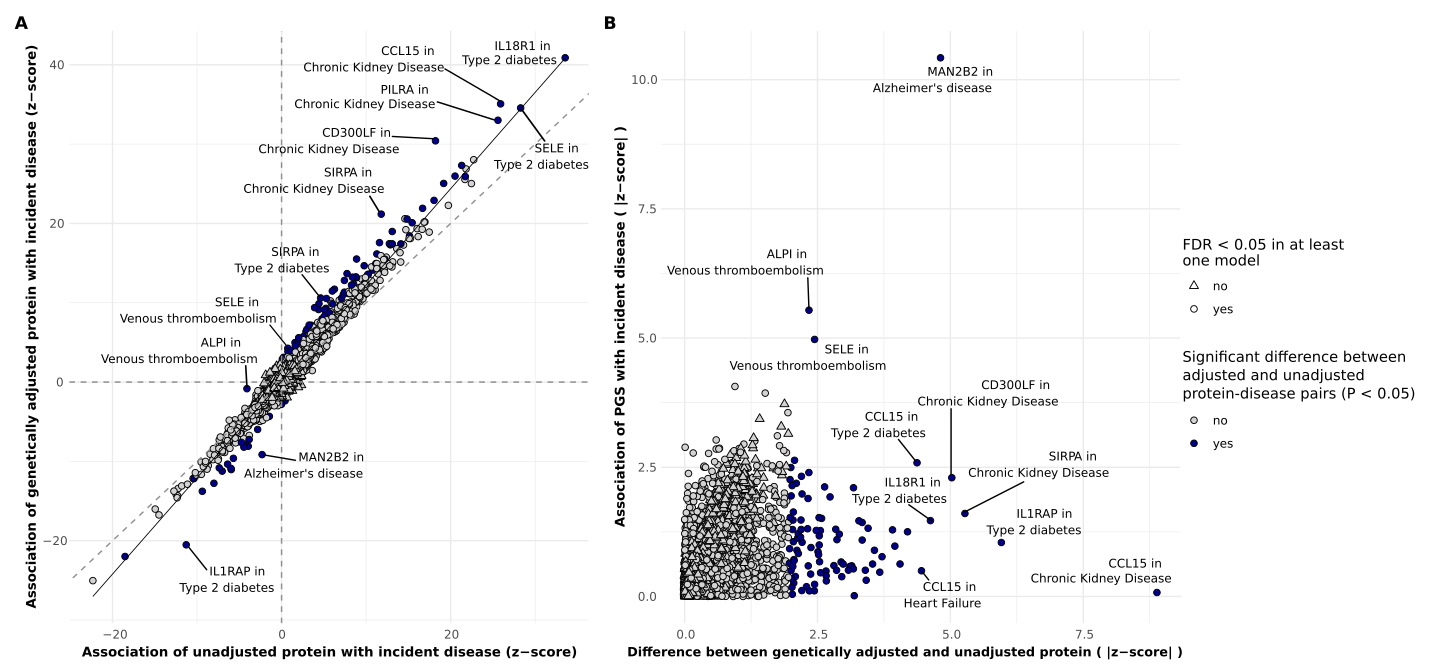

**Supplementary Figure 3 Associations between 94 proteins and 37 diseases in UK Biobank. A.** Logistic regression z-scores for associations with incident diseases for unadjusted proteins (x-axis) and genetically adjusted proteins (y-axis). **B.** Absolute differences in z-score from genetically adjusted and unadjusted proteins models (x-axis) and absolute z-score for the association between protein PGS and diseases (y-axis).
Blue colour indicates a significant difference in effect sizes between adjusted and unadjusted protein associations (p < 0.05), shape indicates FDR significance in at least one model (unadjusted or adjusted protein).

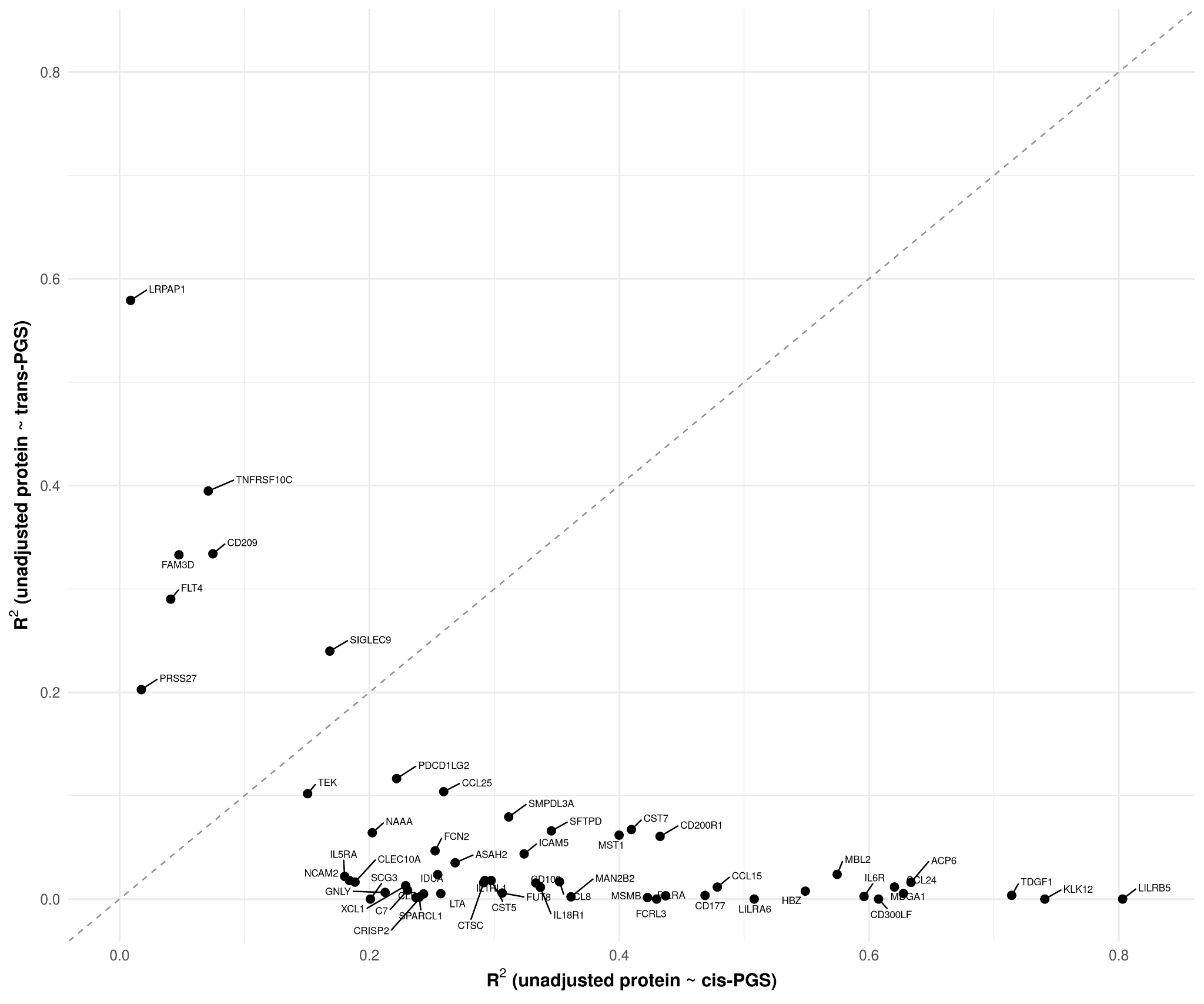

**Supplementary Figure 4 Comparison of protein variance explained by cis- and trans-PGS.** Protein variance explained (R²) by cis-PGS (x-axis) and trans-PGS (y-axis). Each point represents a protein with both cis and trans variants derived from OmicsPred. Proteins with only both cis- or trans- variants are not shown.

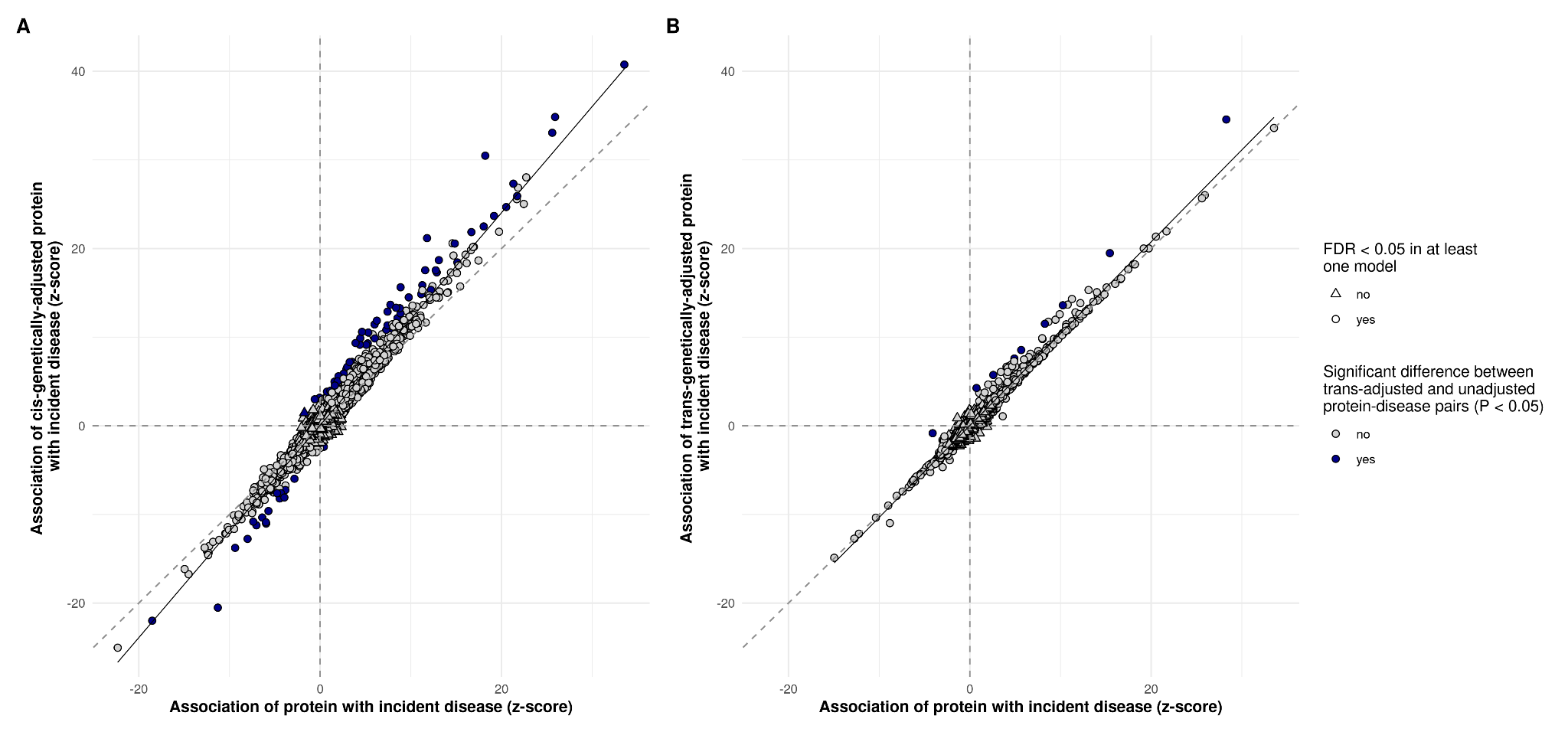

**Supplementary Figure 5 Results from cis- and trans- adjusted logistic regressions.** Logistic regression z-scores for associations of unadjusted proteins (x-axes) and cis-adjusted proteins (A, y-axis) or trans-adjusted proteins (B, y-axis) with incident diseases. Blue colour indicates a significant difference in effect sizes between adjusted and unadjusted protein associations (p < 0.05), and shape indicates FDR significance in at least one model (unadjusted or adjusted protein).

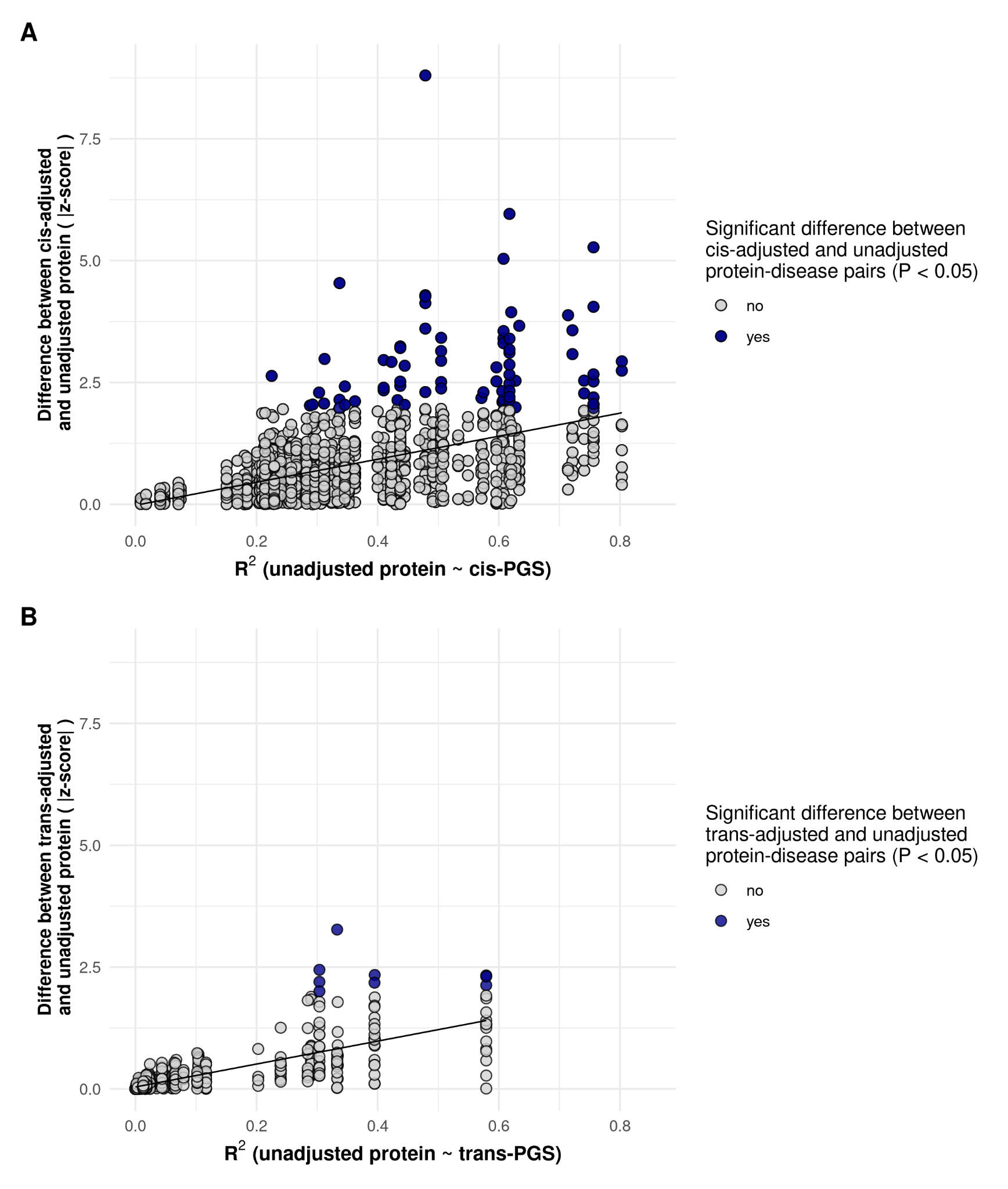

**Supplementary Figure 6 Association between PGS-explained protein variance and cis- or trans- adjusted disease associations.** Relationship between the protein variance explained (R²) by the corresponding PGS and the absolute z-score of the difference between the unadjusted protein and cis- (A) or trans- (B) adjusted protein associations with diseases. We report only significant (FDR < 0.05) protein-disease associations in at least one model (unadjusted or corresponding adjusted protein),
Colour indicates a significant difference in effect sizes between adjusted and unadjusted protein associations (p < 0.05).

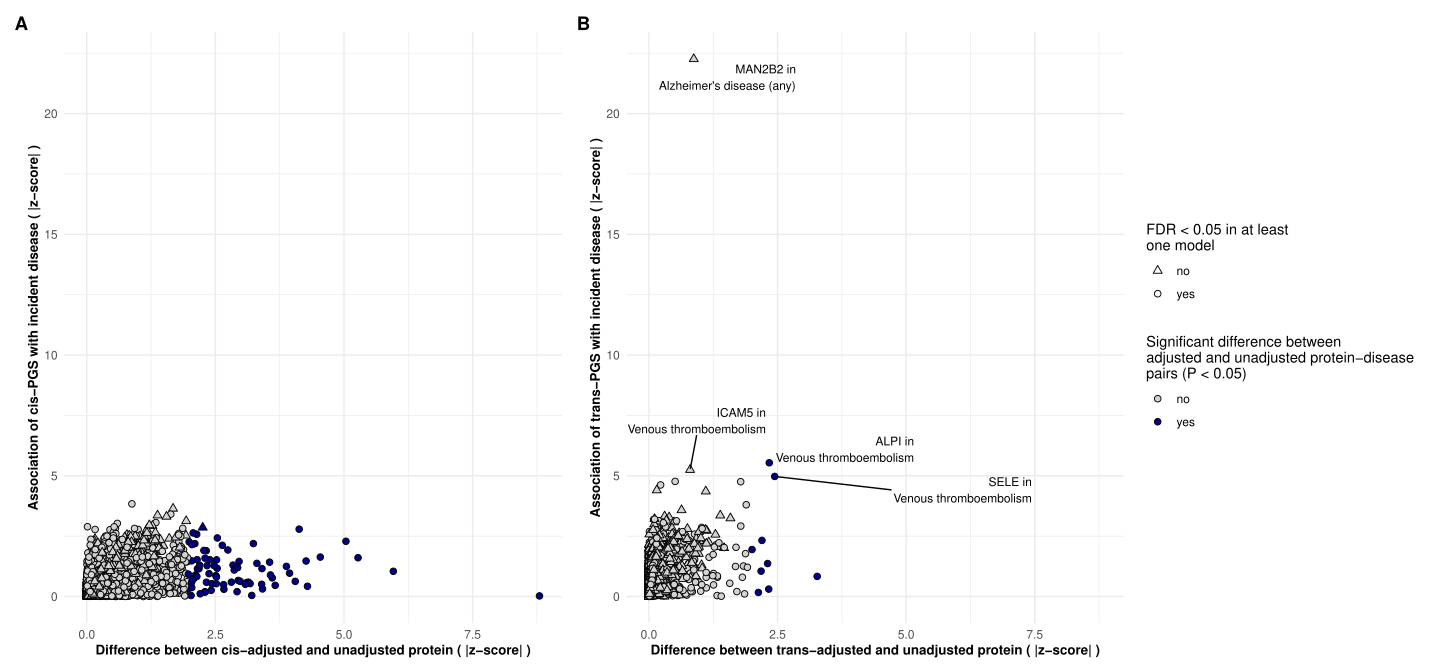

**Supplementary Figure 7 Associations between cis- and trans- PGS with diseases compared to the difference between respective adjusted and unadjusted models.** Absolute differences in z-score from respective cis- (A) or trans- (B) genetically adjusted and unadjusted proteins models (x-axes) and absolute z-score for the association between protein PGS and diseases (y-axes).
Blue colour indicates a significant difference in effect sizes between adjusted and unadjusted protein associations (p < 0.05), shape indicates FDR significance in at least one model (unadjusted or adjusted protein). Labels are present only if PGS–disease association is FDR < 0.05.

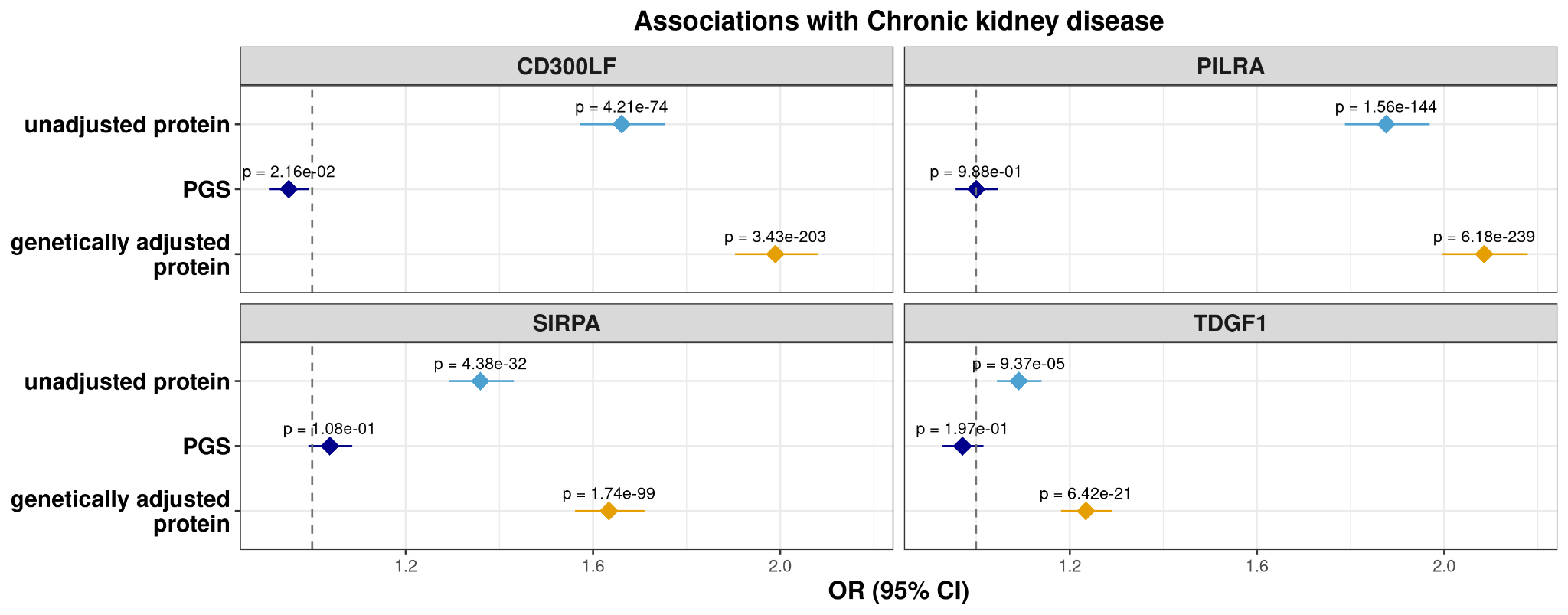

**Supplementary Figure 8 Top associations with chronic kidney disease.**

Logistic regression Odds Ratios (ORs) with 95% confidence intervals are shown for the association of four proteins–CD300LF, PILRA, SIRPA and TDGF1– using unadjusted protein levels, PGS, and genetically adjusted protein, with chronic kidney disease.

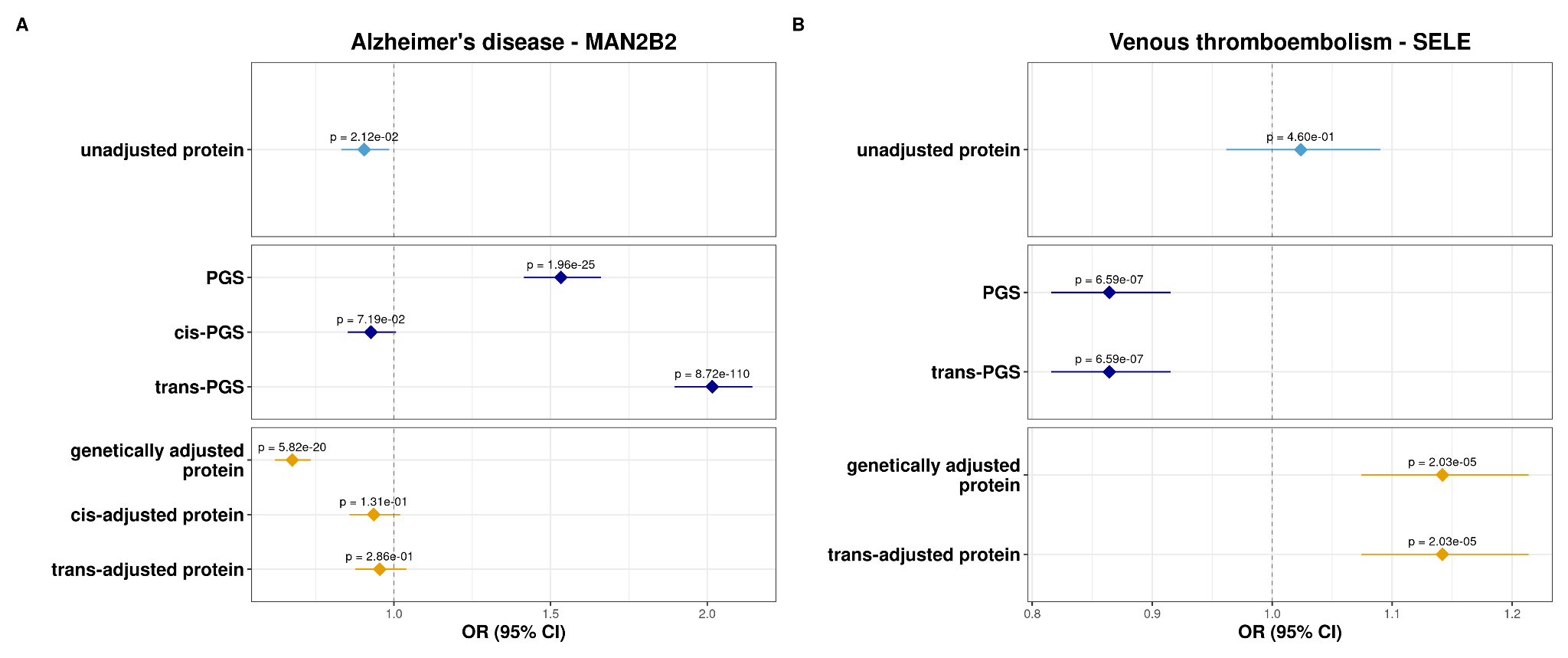

**Supplementary Figure 9 Associations of MAN2B2 and SELE with disease risk.** Logistic regression Odds Ratios (ORs) with 95% confidence intervals are shown for the association MAN2B2 with Alzheimer’s disease (A), and SELE with venous thromboembolism (B) using unadjusted protein, PGS, cis-PGS, trans-PGS, genetically adjusted protein, cis- and trans- adjusted protein. SELE-PGS has been computed with only trans-variants, therefore SELE-cis-PGS is missing.

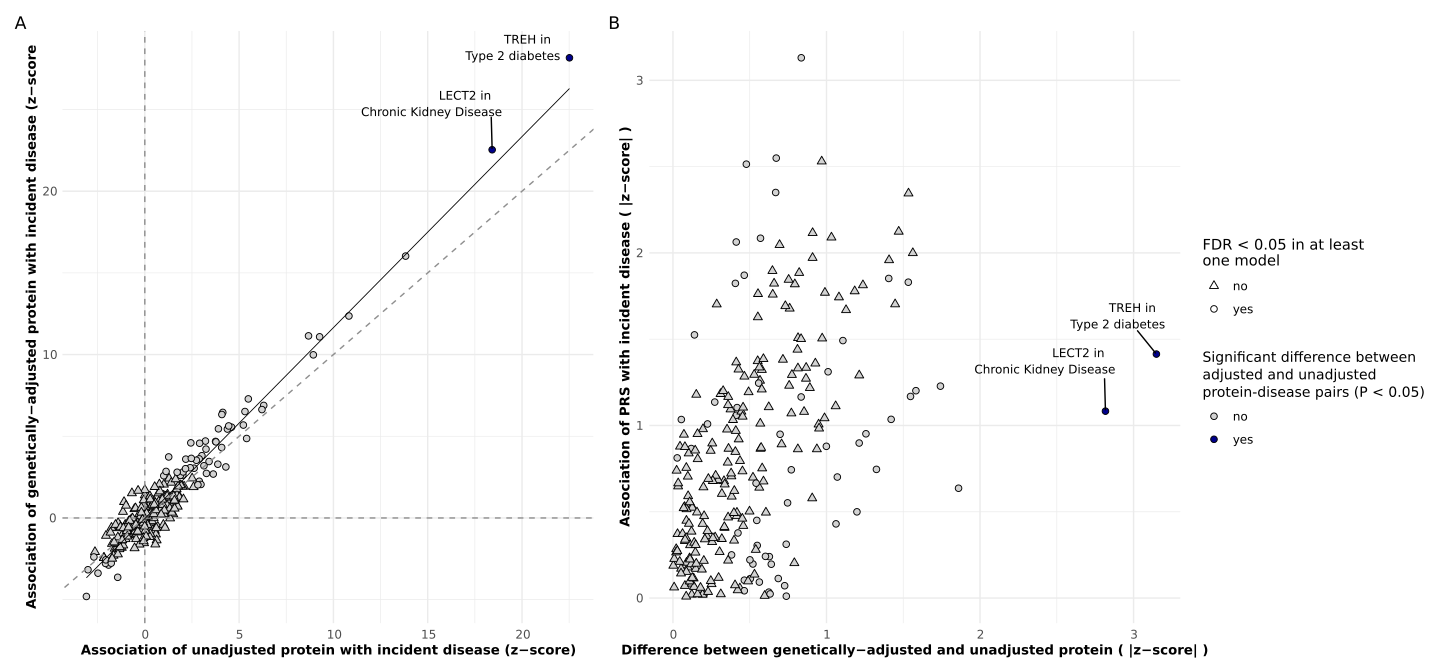

**Supplementary Figure 10 Associations between 7 additional proteins and 37 diseases in UK Biobank, using PGS weights from FinnGen. A.** Logistic regression z-scores for associations with incident diseases for unadjusted proteins (x-axis) and genetically adjusted proteins (y-axis). **B.** Absolute differences in z-score from genetically adjusted and unadjusted proteins models (x-axis) and absolute z-score for the association between protein PGS and diseases (y-axis).
Blue colour indicates a significant difference in effect sizes between adjusted and unadjusted protein associations (p < 0.05), and shape indicates FDR significance in at least one model (unadjusted or adjusted protein).
